# Stringency of the containment measures in response to COVID-19 inversely correlates with the overall disease occurrence over the epidemic wave

**DOI:** 10.1101/2021.01.26.21250501

**Authors:** R. Mezencev, C. Klement

## Abstract

Non-pharmaceutical interventions (NPIs) were the only viable choice to mitigate or suppress transmission of COVID-19 in the absence of efficient and safe vaccines. Moreover, the importance of some NPIs is likely to remain in the future, at least in specific settings, in which the limited vaccination coverage and the high rate of contacts would enable further disease transmission. Nonetheless, the benefits of NPIs have been questioned with respect to their effectiveness and societal costs. In this study of 28 European countries during the first wave of epidemic we demonstrate a significant inverse correlation between the stringency of adopted containment measures and cumulative incidences of the confirmed COVID-19 cases. Our results indicate that early implementation of the stringent containment measures prior to detection of the first confirmed case, and rapid ramp-up of containment stringency after the first case was diagnosed, were instrumental for lowering the number of COVID-19 cases during the epidemic wave. The impact of delayed adoption of containment measures could not be fully attenuated by later adoption of even more stringent community containment.

## 1. Introduction

The continuing pandemic of the coronavirus disease 2019 (COVID-19) is a significant public health concern. Due to its considerable transmission and high level of morbidity and mortality especially among individuals with advanced age and underlying co-morbidities, this disease triggered an unprecedented global “lock-down” in an attempt to control its spread^1^. First emerging in December 2019 as a cluster of pneumonia cases of unknown origin in Wuhan, the capital city of Hubei Province in China, the local outbreak rapidly expanded by travel, nosocomial infection, and close-contact transmission in families. By 23^rd^ January 2020, when strict epidemic control measures were adopted, COVID-19 affected 29 provinces in mainland China and 6 other countries^2^. By 11^th^ March 2020, when the diseases affected 114 countries, the World Health Organization (WHO) declared the rapidly spreading outbreak as pandemic. According to the data compiled by the John Hopkins University Center for Systems Science and Engineering, on 26 January 2021, the cumulative number of confirmed cases exceeded 100 million worldwide, of which more than 2.1 million were fatal^3^.

In the absence of vaccines or chemoprevention, only non-pharmaceutical interventions (NPIs) were available for public health response to the COVID-19 pandemic before December 2020, when vaccination programs started in several countries. These NPIs include: (i) case containment measures targeting individuals through early case detection, contact tracing, isolation of cases and quarantine of contacts, (ii) community containment measures, including various degrees of travel restrictions and social distancing measures, (iii) infection control measures, such as hand hygiene, respiratory etiquette, environmental cleaning, and the use of respiratory protection or face coverings, and (iv) public education.

Imposition of unprecedented containment measures in China, which included cordon sanitaire set up in Hubei Province, aroused controversies regarding their efficacy and societal costs. Along these lines, on 29^th^ February 2020 the WHO advised against travel and trade restrictions to countries experiencing COVID-19 outbreaks^4^. This position reflected the purpose of the WHO International Health Regulations, which is to “*prevent, protect against, control and provide a public health response to the international spread of disease in ways that are commensurate with and restricted to public health risks, and which avoid unnecessary interference with international traffic and trade*”^5^.

Resistance to the implementation of some community containment measures stemmed from concerns, which were previously raised about the effectiveness of the NPIs in control of some epidemics. For instance, discussions about the response to an influenza H5N1 pandemic revealed doubts about the existence of adequate scientific support for some severe social distancing measures^6^. Similarly, the effects of NPIs on 1918-1919 influenza H1N1 pandemic were found to be transient at best, and the cordon sanitaire set up in Liberia during the 2013-2016 Ebola epidemic was found counterproductive and potentially increasing the risk of disease transmission^7^. These controversies may have contributed to the reluctance and delays in the adoption of travel restrictions, and possibly some other community containment measures in response to the COVID-19 pandemic. For this reason, evaluation of effectiveness and socioeconomic impact of the NPIs is needed to inform the epidemic risk management.

In this study, we examined the association between stringency of containment and cumulative incidence of the COVID-19 cases in the first wave of pandemic across 28 European countries. Europe became an epicenter of pandemic early as the disease spread cross-borders both globally and regionally, which led to the restrictions on the entry to the USA for travellers from 26 European countries from Schengen Area starting on 11 March 2020. Nevertheless, European countries displayed remarkable variations in the disease occurrence^8^, which allows studying the role of differences in stringency of containment measures across various European countries and possible identification of patterns responsible for better epidemic control.

### 2. Datasets and Methods

This study considered 28 European countries: 25 EU countries (AT, BE, BG, HR, CY, CZ, DK, EE, FI, FR, DE, HU, IE, IT, LV, LT, LU, NL, PL, PT, RO, SK, SI, ES, SE), two EFTA countries (CH, NO) and the UK (country codes explained in Table 1). For each of these countries, population estimates for 2020 were retrieved from the world statistic project “Worldometer”^9^

**Table 1.** Demographic and epidemiological characteristics of 28 European countries

| Country (code) | Day 1 | First Day of CHI measures<sup>i</sup> | Duration (days)<sup>ii</sup> | Number of cases | Population (2020) | Area (km<sup>2</sup>) | Population Density (per km<sup>2</sup>) | Cumulative Incidence (per 100,000) | Cumulative Incidence Rank |
| --- | --- | --- | --- | --- | --- | --- | --- | --- | --- |
| Austria (AT) | 25-02-20 | 01-02-20 | 103 | 16898 | 9006398 | 83858 | 107.40 | 187.62 | 14 |
| Belgium (BE) | 04-02-20 | 28-01-20 | 144 | 61106 | 11589623 | 30510 | 379.86 | 527.25 | 2 |
| Bulgaria (BG) | 08-03-20 | 03-02-20 | 78 | 2427 | 6948445 | 110994 | 62.60 | 34.93 | 27 |
| Croatia (HR) | 25-02-20 | 27-01-20 | 98 | 2247 | 4105367 | 56594 | 72.54 | 54.73 | 25 |
| Cyprus (CY) | 09-03-20 | 05-03-20 | 109 | 992 | 1207359 | 9251 | 130.51 | 82.16 | 21 |
| Czechia (CZ) | 01-03-20 | 24-01-20 | 94 | 9364 | 10708981 | 78866 | 135.79 | 87.44 | 20 |
| Denmark (DK) | 27-02-20 | 27-02-20 | 132 | 12888 | 5792202 | 44493 | 130.18 | 222.51 | 12 |
| Estonia (EE) | 27-02-20 | 12-03-20 | 122 | 1986 | 1326535 | 45339 | 29.26 | 149.71 | 16 |
| Finland (FI) | 29-01-20 | 27-01-20 | 158 | 7248 | 5540720 | 338145 | 16.39 | 130.81 | 17 |
| France (FR) | 24-01-20 | 23-01-20 | 148 | 191304 | 65273511 | 551695 | 118.31 | 293.08 | 9 |
| Germany (DE) | 27-01-20 | 22-01-20 | 132 | 185450 | 83783942 | 357386 | 234.44 | 221.34 | 13 |

<sup>i</sup> The date (after the 1<sup>st</sup> January 2020) when the Containment and Health Index first exceeded 0
<sup>ii</sup> Duration of the first epidemic wave

| Country (code) | Day 1 | First Day of CHI measures<sup>i</sup> | Duration (days)<sup>ii</sup> | Number of cases | Population (2020) | Area (km<sup>2</sup>) | Population Density (per km<sup>2</sup>) | Cumulative Incidence (per 100,000) | Cumulative Incidence Rank |
| --- | --- | --- | --- | --- | --- | --- | --- | --- | --- |
| Hungary (HU) | 04-03-20 | 28-01-20 | 114 | 4123 | 9660351 | 93030 | 103.84 | 42.68 | 26 |
| Ireland (IE) | 29-02-20 | 04-02-20 | 119 | 25414 | 4937786 | 70273 | 70.27 | 514.68 | 4 |
| Italy (IT) | 31-01-20 | 23-01-20 | 160 | 242149 | 60461826 | 301338 | 200.64 | 400.50 | 6 |
| Latvia (LV) | 02-03-20 | 29-01-20 | 112 | 1111 | 1886198 | 64589 | 29.20 | 58.90 | 24 |
| Lithuania (LT) | 28-02-20 | 29-01-20 | 122 | 1815 | 2722289 | 65300 | 41.69 | 66.67 | 23 |
| Luxembourg (LU) | 29-02-20 | 01-03-20 | 97 | 4027 | 625978 | 2586 | 242.06 | 643.31 | 1 |
| Netherlands (NL) | 27-02-20 | 27-01-20 | 128 | 50487 | 17134872 | 41198 | 415.92 | 294.64 | 8 |
| Norway (NO) | 26-02-20 | 31-01-20 | 138 | 8981 | 5421241 | 385178 | 14.07 | 165.66 | 15 |
| Poland (PL) | 04-03-20 | 23-01-20 | 122 | 35405 | 37846611 | 312685 | 121.04 | 93.55 | 19 |
| Portugal (PT) | 02-03-20 | 26-01-20 | 77 | 29036 | 10196709 | 91568 | 111.36 | 284.76 | 10 |
| Romania (RO) | 26-02-20 | 27-01-20 | 93 | 18791 | 19237691 | 238397 | 80.70 | 97.68 | 18 |
| Slovakia (SK) | 06-03-20 | 01-01-20 | 88 | 1522 | 5459642 | 49036 | 111.34 | 27.88 | 28 |
| Slovenia (SI) | 05-03-20 | 04-03-20 | 82 | 1469 | 2078938 | 20273 | 102.55 | 70.66 | 22 |
| Spain (ES) | 01-02-20 | 24-01-20 | 138 | 244683 | 46754778 | 498511 | 93.79 | 523.33 | 3 |
| Sweden (SE) | 01-02-20 | 31-01-20 | 101 | 27301 | 10099265 | 450295 | 22.43 | 270.33 | 11 |
| Switzerland (CH) | 25-02-20 | 15-02-20 | 99 | 30874 | 8654622 | 41290 | 209.61 | 356.73 | 7 |
| United Kingdom (UK) | 31-01-20 | 20-01-20 | 159 | 286349 | 67886011 | 242495 | 279.95 | 421.81 | 5 |

Cumulative numbers of the confirmed COVID-19 cases were downloaded on 13^th^ September 2020 as time series from the COVID-19 Data Repository (Center for Systems Science and Engineering (CSSE), Johns Hopkins University)^3^. The dataset covered period from 22^nd^ January to 12^th^ September 2020. Cumulative numbers per day were presented as scatterplots starting from the day of the first confirmed case of COVID-19 (day 1) per each country. For each curve of cumulative numbers of COVID-19 cases, first and second derivative curves were plotted based on the numerical differentiation and smoothing by the Lowess method (medium, 10 points in smoothing window) using GraphPad Prism version 8.0.1.244 for Windows (GraphPad Software, San Diego, California USA). Scatterplots were used for determination of cumulative incidence (CI) and the end day of the first epidemic wave in each country.

For the purpose of this report, epidemic wave is considered as a sigmoidal curve of cumulative cases with four distinguishable stages: (i) lagging phase with marginal daily increase in case numbers, (ii) acceleration stage with increasing number of daily cases, (iii) deceleration stage with decreasing number of new cases, and (v) stationary stage with marginal daily increases and stagnation of total number of cases. The first order derivative curve (growth rate graph) is approximately bell shaped and the second order derivative (growth acceleration) consists of two bell-shaped curves^10^ (these patterns are shown on the Supplemental figure 1). The end day of the first wave and the cumulative incidence for the first wave of epidemic in each country were identified by examining patterns of these three curves, allowing for transient stationary intervals after identifiable peaks in first order derivative curves.

**Figure 1.**
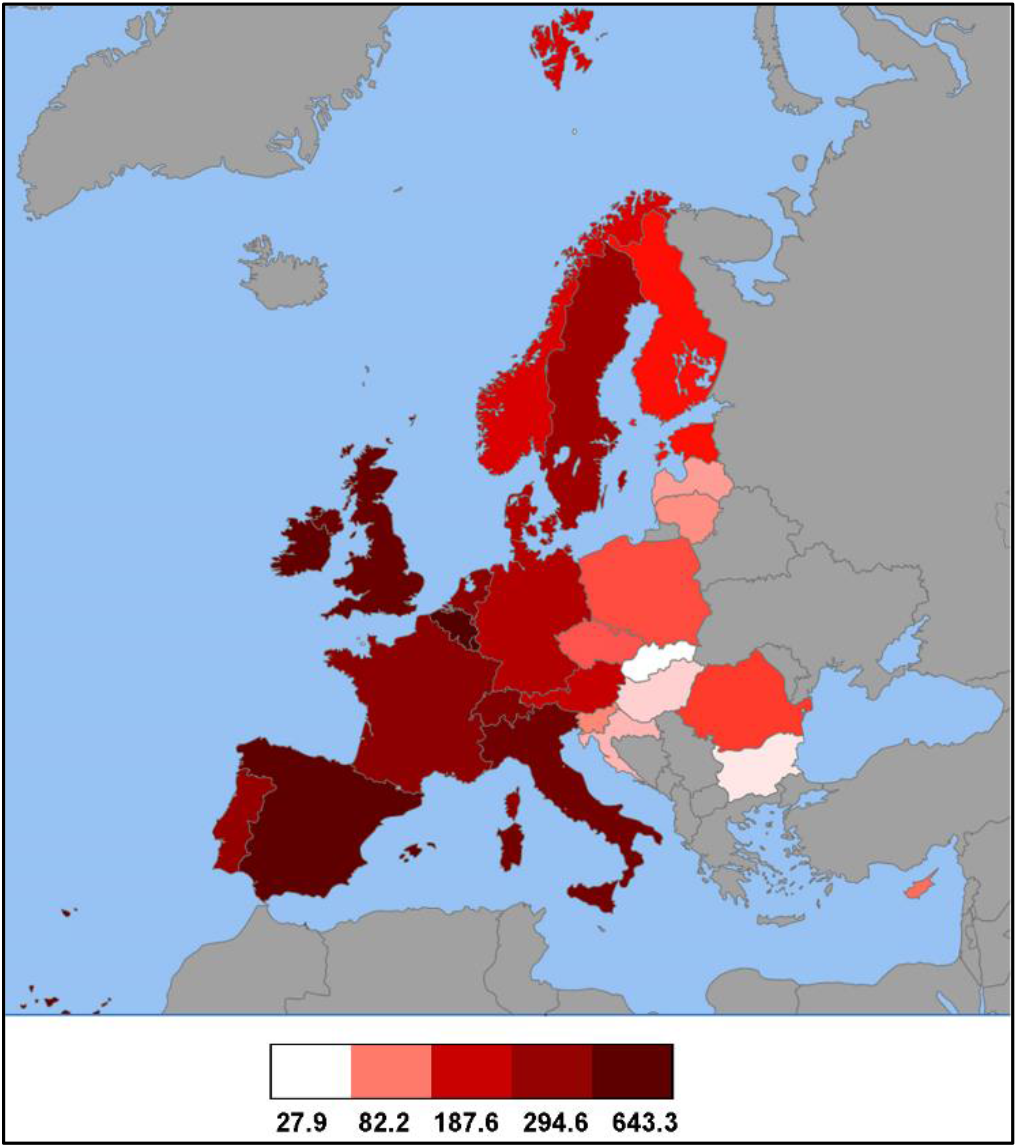
Cumulative incidence of COVID-19 cases in 28 European countries at the end of the first epidemic wave. Color coding reflects the number of confirmed cases per 100,000 people. Gray color - data not shown.

Containment and Health Index (CHI) is one of the four aggregate indices reported by the Oxford COVID-19 Government Response Tracker (OxCGRT) project from the Blavatnik School of Government.^11^ These indices are calculated from indicators on (i) containment and closure policies (C1-C8), (ii) economic policies (E1-E4), and (iii) health system policies (H1-H7). Containment and Health Index (CHI) is composed of the following 11 individual containment and health response indicators recorded on ordinal scales: School closing (C1), Workplace closing (C2), Public events cancellation (C3), Restriction on gathering size (C4), Public transportation closing (C5), Stay at home orders (C6), Restrictions on internal movement (C7), Restriction on international travel (C8), Public information campaign (H1), Testing policy (H2), and Contact tracing (H3).^11^ CHI values (a “display” version) for all EU countries and for each day of the first wave of COVID-19 epidemic were downloaded as an “OxCGRT_latest.csv” file on 30 September 2020.^12^ Cumulative CHI indices (cCHIs) from the day 1 of epidemic were determined for each Day D as the sum of CHIs for all days starting with day of the first confirmed diagnosis of COVID-19 (day 1) up to the Day D. Cumulative CHI indices for pre-epidemic period in each country (cCHI(<1)) were determined by summing CHI values from 01 January 2020 to the day preceding day 1 of epidemic in each country.

Degree of association between cumulative incidences and cumulative CHI values was determined using Spearman’s semi-partial correlations by eliminating the effect of population density on cumulative incidence, using the package “ppcor”^13^ in R Environment version 3.5.1 (R Core Team, Vienna, Austria; https://www.R-project.org). Two-sided p-values for significance of the Spearman’s correlation were adjusted using the Benjamini-Hochberg procedure implemented in p.adjust function in R Environment. Zero-order correlations between two variables without controlling for the influence of other variables were determined as Spearman’s rank-order correlations using GraphPad Prism version 8.0.1.244 for Windows. All reported p-values are two-tailed. Hierarchical clustering (Euclidian distance, average linkage) was performed on cCHI values using the CIMiner tool (http://discover.nci.nih.gov/cimminer).

Apple Mobility Trends Reports were accessed as the complete data in the .csv file format on 22^nd^ December 2020^14^.The data reflect the number of requests for directions in “Apple Maps” relative to the baseline on 13 January 2020, when each component of mobility is assigned the value of 100%). The driving, walking and transit transportation data were extracted for selected countries on a country level for each day after 13^th^ January 2020 and their centered 7-day averages were calculated for each day and plotted over time.

Google Community Mobility Reports were accessed on 13^th^ December 2020. These data include mobility trends in six different categories (“Grocery and Pharmacy”, “Parks”, “Transit Stations”, “Workplaces”, and “Residential Places”) determined based on the location history for a sample of Google accounts and expressed relative to the baseline. The baseline is the median value for the corresponding day of the week across 5-week period from 3 January to 6 February 2020.^15^ For this study, only mobility trends for places of residence were used, which represent duration of the time spent at places of residence relative to the baseline.

## 3. Results

### 3.1 Occurrence of COVID-19 over the first epidemic wave differed considerably across selected European countries

The last days of the first epidemic waves of COVID-19 in 28 European countries, and corresponding cumulative numbers of confirmed cases (Table 1) were determined from the scatterplots of cumulative numbers of cases vs. days, and their first and second order derivative curves (Supplemental figures 2-6).

**Figure 2.**
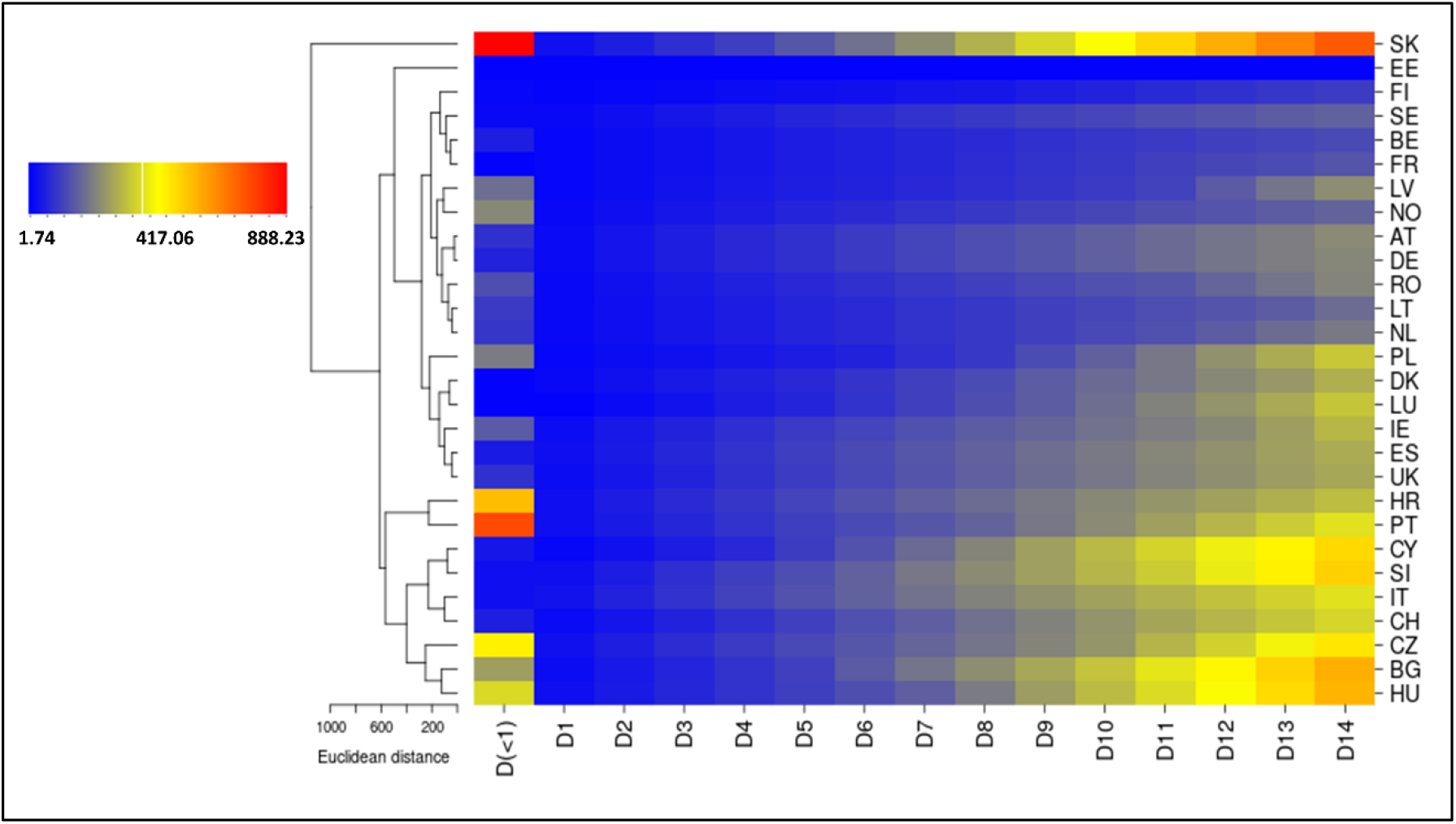
Hierarchical clustering of cumulative Containment and Health Indices (cCHIs) for pre-epidemic period (D<1) and epidemic days 1-14 across 28 countries. Distance method: Euclidian; Cluster algorithm: Average linkage.

The first waves of the COVID-19 epidemics in these countries started between 24^th^ January and 9^th^ March 2020, and lasted for 77-160 days (median 116.5 days). Cumulative incidence of confirmed cases displays high variability ranging 27.88-643.31 cases per 100,000 population (Table 1, Figure 1), and appears to have a multimodal distribution (Supplemental figure 7).

Cumulative incidence is positively and statistically significantly correlated with population density (Spearman’s ρ=0.467; CI_95_:0.102-0.721; p-value=0.0123; Supplemental figure 8A) and with the duration of the first epidemic wave (Spearman’s ρ=0.460; CI95:0.093-0.717; p-value=0.0138; Supplemental figure 8B). In addition, the cumulative incidence remains positively and significantly correlated with population density while controlling cumulative incidence for the effect of epidemic duration (semi-partial Spearman’s ρ=0.450, p-value=0.0185, test statistic=2.521). These results indicate that the overall risk of COVID-19 over the first epidemic waves in European countries increased with increasing population density.

Based on the cumulative incidence of confirmed COVID-19 cases, three highest ranking countries were identified as Luxembourg, Belgium and Spain. In contrast, Slovakia, Bulgaria and Hungary reached the lowest cumulative incidences over the first epidemic waves (Figure 1 and Table 1).

### 3.2 Stringency of the containment measures differed across European countries relative to the beginning of their COVID-19 epidemics

Containment and health measures that are reflected in the CHI Containment and Health Indices (CHI) were first adopted in 28 European countries between 1^st^ January and 12^th^ March 2020 (Table 1). Intriguingly, Slovakia was the only country among 28 considered European countries, and one of 10 countries globally, which had some containment and health measures in place already on 01 January 2020. The CHI values accumulated over pre-epidemic period (cCHI(<Day1)) show considerable differences across individual European countries (Table 2). This measure, which reflects stringency of the pre-epidemic containment measures, was highest in Slovakia (889.97), and lowest in Denmark, Estonia and Luxembourg (0). Similarly, considerable differences across European countries were found for cumulative CHI values accumulated from day 1 up to the day 14 (Table 2), indicating different stringency of containment measures adopted in individual countries from the first days of their epidemics. Per this metrics, Slovakia ranks first among 28 European countries for days 5-14, and second for days 1-4 from the beginning of her epidemic (Table 2). Unsupervised hierarchical clustering for cumulative stringencies across days 1 to 14 with and without inclusion of cumulative stringencies for pre-epidemic periods identified clusters of countries with similar stringencies of adopted containment measures (Figures 2 and 3). Interestingly, Slovakia forms a singleton in both cluster analyses, indicating its unique status (outlier) among European countries due to the high stringency and early timing of the containment measures adopted in that country. Considerable similarity is also visible for the low-cumulative incidence countries Bulgaria and Hungary, as well as Slovenia and Cyprus (Figure 3). Bulgaria and Hungary show an appreciable stringency of CHIs in the pre-epidemic period, as well as early ramping-up of their stringencies after the diagnosis of first COVID-19 cases, while Slovenia and Cyprus had considerably lower pre-epidemic stringency of containment, but rapidly increasing cCHIs from the first days of their epidemics (Figure 2 and Table 2). On the other hand, high-incidence countries display a more complex pattern of stringency over time, exemplified by differences between similar pairs BE and FR versus UK and ES (Figures 2 and 3).

**Table 2.** Cumulative values of the Containment and Health Index (cCHI) accumulated over pre-epidemic priods (<Day 1) or between day 1 of epidemic and day 2-14 (D2-D14). Color coding reflects the value of cCHI (for pre-epidemic period: white=low, green=high; for epidemic period: blue=low, red=high).

| Day | <Day 1 | D1 | D2 | D3 | D4 | D5 | D6 | D7 | D8 | D9 | D10 | D11 | D12 | D13 | D14 |
| --- | --- | --- | --- | --- | --- | --- | --- | --- | --- | --- | --- | --- | --- | --- | --- |
| AT | 81.81 | 16.67 | 33.34 | 50.01 | 66.68 | 83.35 | 100.02 | 116.69 | 133.36 | 150.03 | 166.7 | 183.37 | 200.04 | 216.71 | 240.19 |
| BE | 50.01 | 9.09 | 18.18 | 27.27 | 36.36 | 45.45 | 54.54 | 63.63 | 72.72 | 81.81 | 90.9 | 99.99 | 109.08 | 118.17 | 127.26 |
| BG | 272.68 | 20.45 | 40.9 | 61.35 | 86.35 | 111.35 | 156.05 | 200.75 | 245.45 | 290.15 | 339.39 | 400 | 460.61 | 521.22 | 584.1 |
| HR | 559.76 | 23.48 | 46.96 | 70.44 | 93.92 | 117.4 | 140.88 | 164.36 | 187.84 | 211.32 | 234.8 | 258.28 | 281.76 | 305.24 | 328.72 |
| CY | 36.36 | 9.09 | 27.27 | 45.45 | 68.18 | 104.54 | 140.9 | 181.81 | 228.78 | 275.75 | 322.72 | 369.69 | 416.66 | 463.63 | 510.6 |
| CZ | 468.3 | 25 | 50 | 75 | 100 | 125 | 150 | 175 | 200 | 228.03 | 260.61 | 311.37 | 364.4 | 423.49 | 485.61 |
| DK | 0 | 13.64 | 27.28 | 40.92 | 54.56 | 68.2 | 89.41 | 110.62 | 131.83 | 157.59 | 183.35 | 209.11 | 234.87 | 265.17 | 303.81 |
| EE | 0 | 0 | 0 | 0 | 0 | 0 | 0 | 0 | 0 | 0 | 0 | 0 | 0 | 0 | 0 |
| FI | 9.1 | 4.55 | 9.1 | 13.65 | 18.2 | 22.75 | 27.3 | 31.85 | 36.4 | 47.76 | 59.12 | 70.48 | 81.84 | 93.2 | 104.56 |
| FR | 2.27 | 9.09 | 18.18 | 27.27 | 36.36 | 45.45 | 54.54 | 63.63 | 74.99 | 86.35 | 97.71 | 109.07 | 120.43 | 131.79 | 143.15 |
| DE | 59.1 | 16.67 | 33.34 | 50.01 | 66.68 | 83.35 | 100.02 | 116.69 | 133.36 | 150.03 | 166.7 | 183.37 | 200.04 | 216.71 | 233.38 |
| HU | 378.76 | 21.21 | 42.42 | 63.63 | 84.84 | 106.05 | 134.08 | 162.11 | 215.14 | 271.2 | 327.26 | 383.32 | 439.38 | 506.8 | 574.22 |
| IE | 156.15 | 19.7 | 39.4 | 59.1 | 78.8 | 98.5 | 118.2 | 137.9 | 157.6 | 177.3 | 197 | 216.7 | 236.4 | 272.76 | 318.21 |
| IT | 22.71 | 28.03 | 56.06 | 84.09 | 112.12 | 140.15 | 168.18 | 196.21 | 224.24 | 252.27 | 280.3 | 308.33 | 336.36 | 364.39 | 392.42 |
| LV | 188.56 | 9.85 | 19.7 | 29.55 | 39.4 | 49.25 | 59.1 | 68.95 | 78.8 | 88.65 | 100.77 | 112.89 | 155.31 | 200.01 | 244.71 |
| LT | 100 | 12.12 | 24.24 | 36.36 | 48.48 | 60.6 | 72.72 | 84.84 | 96.96 | 109.08 | 121.2 | 133.32 | 145.44 | 157.56 | 185.59 |
| LU | 0 | 0 | 15.15 | 30.3 | 45.45 | 60.6 | 84.84 | 109.08 | 133.32 | 157.56 | 190.89 | 224.22 | 257.55 | 295.43 | 342.4 |
| NL | 93.93 | 12.12 | 24.24 | 36.36 | 48.48 | 60.6 | 72.72 | 84.84 | 96.96 | 110.6 | 124.24 | 137.88 | 158.33 | 185.6 | 212.87 |
| NO | 236.34 | 12.12 | 24.24 | 36.36 | 48.48 | 60.6 | 72.72 | 84.84 | 96.96 | 109.08 | 121.2 | 133.32 | 145.44 | 157.56 | 172.71 |
| PL | 213.79 | 9.09 | 18.18 | 27.27 | 36.36 | 45.45 | 56.81 | 77.26 | 97.71 | 131.8 | 165.89 | 206.04 | 253.01 | 299.98 | 346.95 |
| PT | 763.56 | 21.21 | 42.42 | 63.63 | 84.84 | 106.05 | 127.26 | 148.47 | 174.23 | 206.81 | 239.39 | 278.03 | 316.67 | 355.31 | 393.95 |
| RO | 134.19 | 13.64 | 27.28 | 40.92 | 54.56 | 68.2 | 81.84 | 95.48 | 109.12 | 122.76 | 136.4 | 150.04 | 175.04 | 202.31 | 229.58 |
| SK | 889.97 | 25.76 | 51.52 | 77.28 | 107.58 | 150 | 192.42 | 244.69 | 307.57 | 374.99 | 442.41 | 515.89 | 589.37 | 662.85 | 736.33 |
| SI | 24.24 | 24.24 | 48.48 | 77.27 | 106.06 | 134.85 | 170.46 | 206.07 | 241.68 | 277.29 | 315.93 | 354.57 | 411.39 | 468.21 | 525.03 |
| ES | 42.42 | 21.21 | 42.42 | 63.63 | 84.84 | 106.05 | 127.26 | 148.47 | 169.68 | 190.89 | 212.1 | 233.31 | 254.52 | 275.73 | 296.94 |
| SE | 12.12 | 12.12 | 24.24 | 36.36 | 48.48 | 60.6 | 72.72 | 84.84 | 96.96 | 109.08 | 121.2 | 133.32 | 145.44 | 157.56 | 169.68 |
| CH | 51.56 | 17.42 | 34.84 | 56.81 | 83.33 | 109.85 | 136.37 | 162.89 | 193.95 | 225.01 | 256.07 | 287.13 | 315.16 | 343.19 | 371.22 |
| UK | 83.38 | 18.94 | 37.88 | 59.09 | 80.3 | 101.51 | 122.72 | 143.93 | 165.14 | 186.35 | 207.56 | 228.77 | 249.98 | 271.19 | 292.4 |

**Figure 3.**
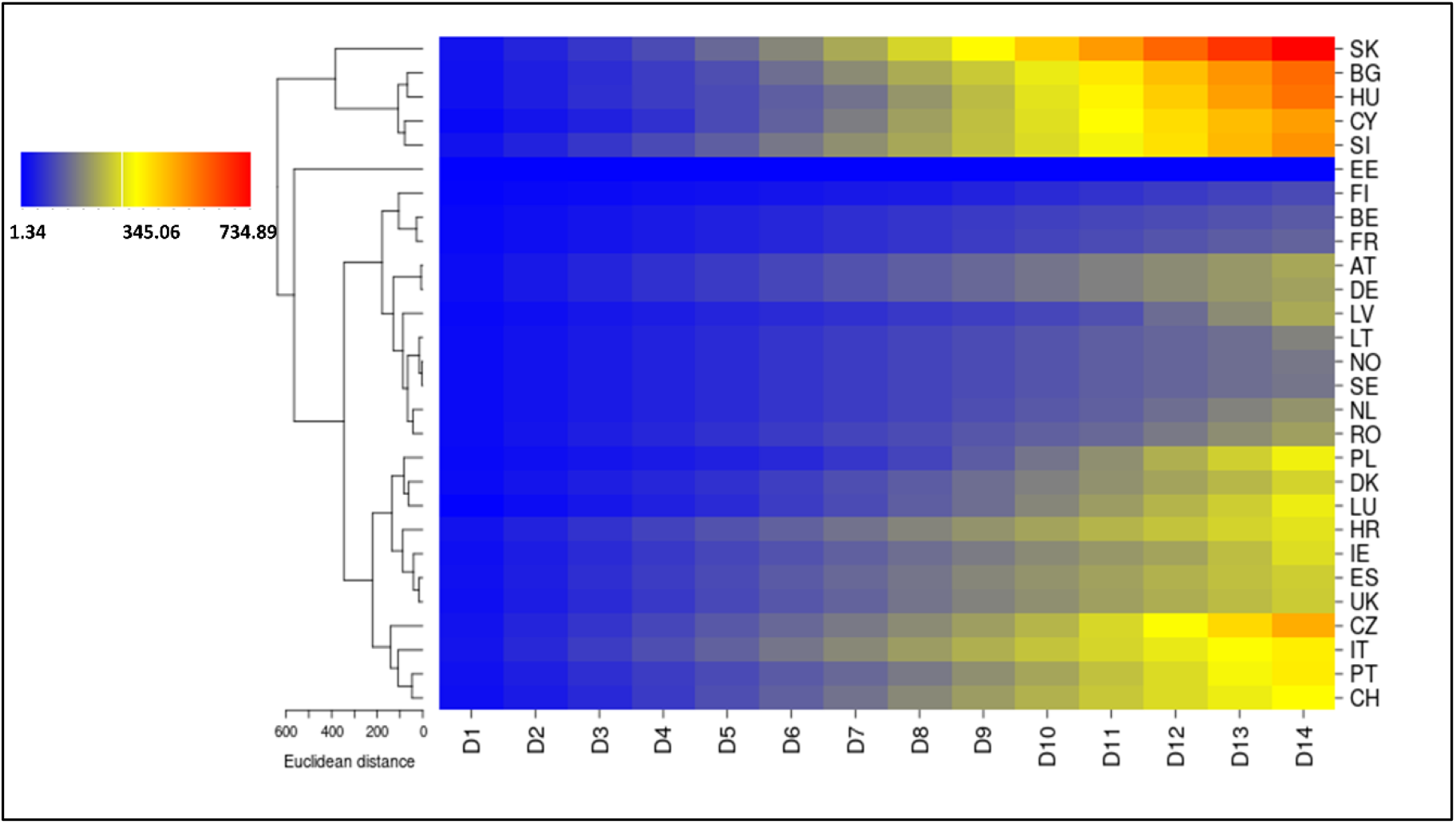
Hierarchical clustering of cumulative Containment and Health Indices (cCHIs) for epidemic days 1-14 across 28 countries. Distance method: Euclidian; Cluster algorithm: Average linkage.

### 3.3 Stringency of the containment measures inversely correlates with disease occurrence across European countries

Slovakia, which displays highest stringency of the pre-epidemic containment measures, as well as the first or the second highest stringency from day 1 of the epidemic, reached the lowest cumulative incidence of the confirmed COVID-19 cases across European countries at the end of the first epidemic wave (Figure 1 and Table 1). Conversely, Luxembourg, with the highest cumulative incidence of cases, displayed the lowest cumulative CHI for period between 1^st^ January and the first day of epidemic. In addition, Luxembourg scored low in the cumulative CHIs from the day 1 of the epidemic, and its cCHI values were lower than median cumulative CHIs for the first 7 days of the epidemics across 28 countries. These findings suggested existing association between cumulative incidence for the first epidemic wave and stringency of the containment measures across European countries (Figure 4).

**Figure 4.**
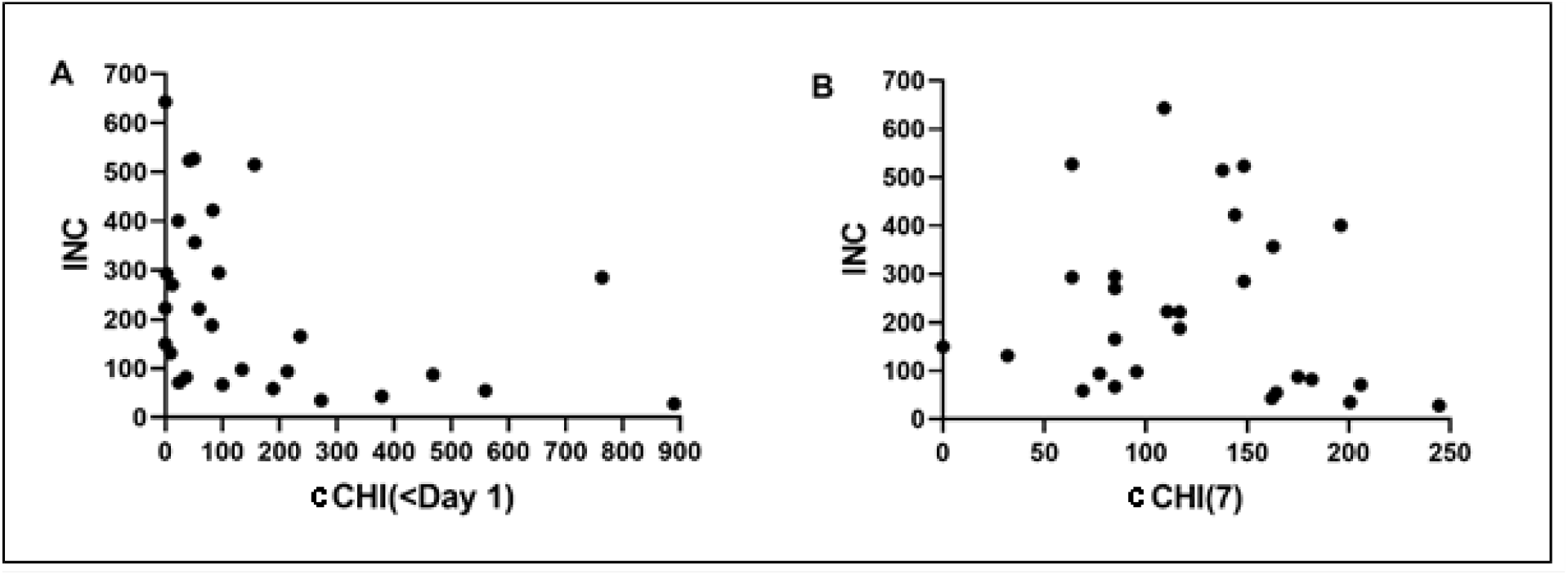
Scatterplot of cumulative incidences of confirmed COVID-19 cases versus: (A) cumulative Containment Health Index prior the first day of epidemic, and (B) cumulative Containment Health Index for the first 7 days of epidemic. INC: cumulative incidence; cCHI(<Day 1): cumulative Containment Health Index prior the first day; cCHI(7): cumulative Containment Health Index for the first 7 days

Next, semi-partial correlations were performed to determine the degree of associations between the cCHI values for specific time periods and cumulative incidences of COVID-19 whilst controlling the cumulative incidence for population density (Table 3). There was a moderate, inverse semi-partial correlation (ρ= −0.50) between the cumulative incidence of COVID-19 cases (average±SD: 225.91±173.95 per 100,000) and cCHI accumulated over the pre-epidemic period (176.11±230.08), whilst controlling for population density (130.99±100.98 km^-2^), which was statistically significant (p-adjusted=0.0247). Likewise, a moderate inverse correlation was found between the cumulative incidence and cumulative CHI over the first 7 or more days of epidemic (123.78±56.24 for the first 7 days) whilst controlling for population density (Table 3). To conclude, higher stringency of containment during the pre-epidemic period, and during the first 7 or more days of the epidemic, is associated with lower overall disease occurrence.

**Table 3.**
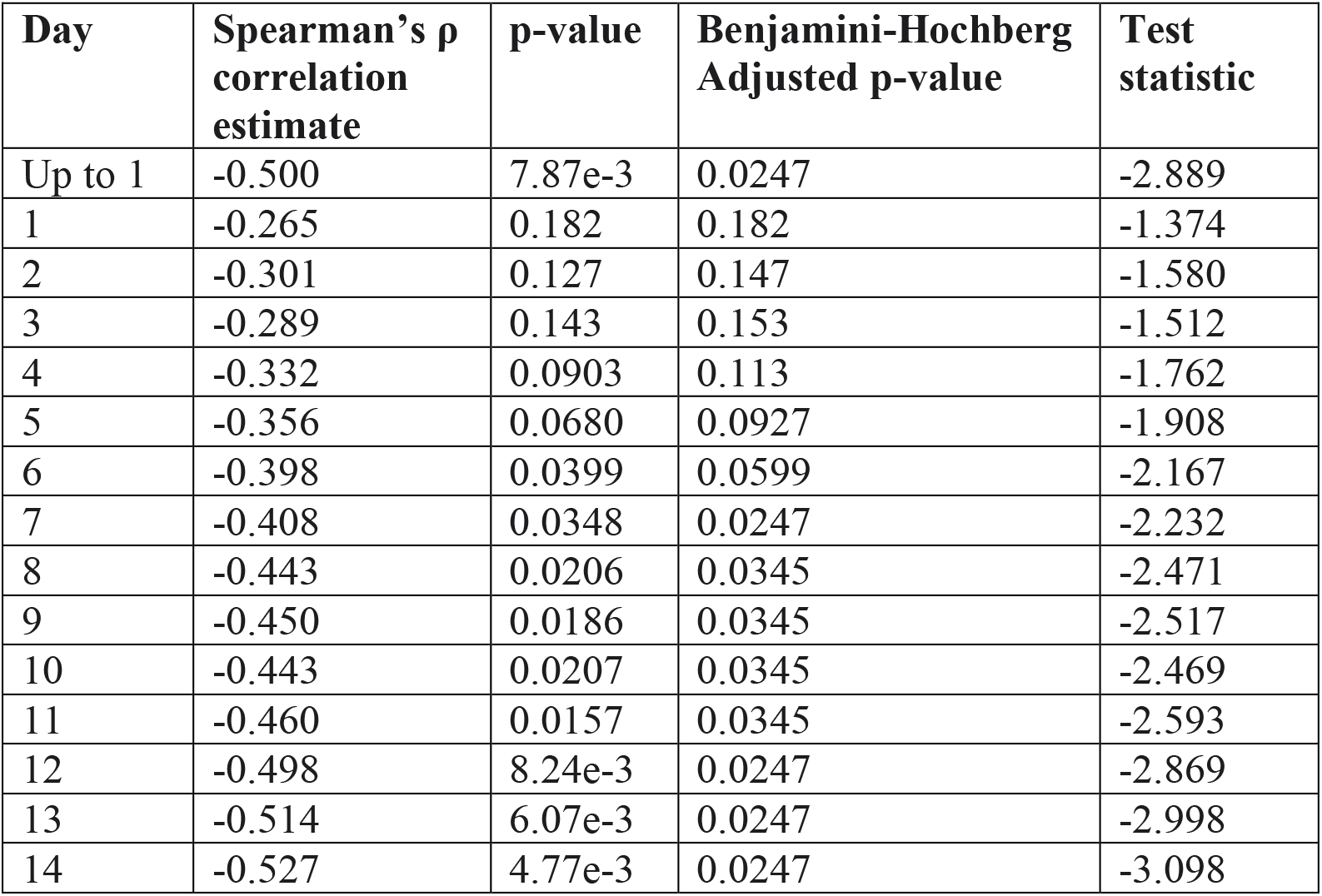
Semi-partial correlations for association between cumulative Containment Health Indices (cCHIs) for various days of epidemic and cumulative incidences of COVID-19 in the first wave

Zero-order correlations showed moderate negative correlation (ρ= −0.50) between the cumulative incidence of COVID-19 and cumulative CHI index over period from 1^st^ January 2020 to the last day before the beginning of epidemic, which was statistically significant (p-value=0.0064, n=28). As a result, population density had very little influence in controlling for the relationship between cumulative incidence of the disease and the pre-epidemic stringency of containment and health measures. In contrast, zero-order correlation between the cumulative incidence and cumulative CHI index over the first 7 days of epidemic was weak and not statistically significant (ρ= −0.26, p=0.18, n=28), and the same applied to the correlation between cumulative CHI index over the first 7 days and population density (ρ= 0.21, p=0.28, n=28). As a result, population density appears to be a suppressor variable that suppressed the effect cCHI on the cumulative incidence.

Interestingly, Portugal featured the second highest pre-epidemic cumulative CHI, but eventually ranked only 10th out of 28 countries with respect to the cumulative COVID-19 incidence. In spite of the relatively stringent pre-epidemic containment measures, that country reached relatively high cumulative incidence, which is consistent with observed lower stringency of the containment measures adopted from the day 1 of the epidemic. The lower stringency of containment from the day 1 of epidemic is demonstrated by Portugal ranking 7th to 10th in the cCHI values for days 1 up to 14. This finding, visualized by an outlier status for Portugal in the scatterplot of cumulative incidences vs. pre-epidemic cCHIs, indicates that pre-epidemic containment measures cannot fully compensate for the lack of adequate stringency of containment once the disease presence is confirmed in the population (Figure 4A, second point from the left).

Countries, which were top ranking based on their cumulative incidences (LU, BE, and ES), typically scored low on the pre-epidemic cCHIs (respective ranks 27, 18, 19), and low or intermediate on the cCHIs for day 14 (ranks 11, 26, 15). In contrast, countries scoring lowest on the cumulative incidence (SK, BG and HU), scored high in the cumulative CHIs both in pre-epidemic period (ranks 1, 6, and 5) and epidemic days 1-14 (ranks 1, 2 and 3). Countries with poor scoring on pre-epidemic cumulative CHIs, such as CY and SI (ranks 20 and 21), could still display relatively low cumulative incidences (ranks 21 and 22), if their cumulative CHIs for day 14 scored high (ranks 5 and 4).

### 3.4 Stringency of the containment measures impacted the mobility of the population

Cumulative CHI values influenced the occurrence of COVID-19 in the first epidemic wave through its impact on mobility, which can be recognized through exploration of the CHI and mobility trends over time. In Slovakia, visualization reveals appreciable inverse relationship between the CHI and mobility (Figure 5). Mobility represented by transit, walking and driving (Apple) tended to decrease, while the time spent in the areas of residence (Google) tended to increase with increasing level of the containment stringency over the first epidemic wave (Figure 3). This association is, however, influenced by compliance with public health measures, which mediates relationship between adopted public health policies and mobility. In addition, this compliance can change over time. The decrease of mobility in Slovakia can be observed for about 3 weeks preceding 6th March 2020, which is the date of the first diagnosis of a confirmed COVID-19 case (day 1). Thereafter, an additional steep decrease in mobility was visible starting from the day 1, which is consistent with increasing values of the CHIs. The mobility reached minimum at about day 18, at which point it started increasing gradually without parallel decrease in the CHI. Increasing mobility without decreasing CHI is suggestive of decreased public compliance up to the day ∼37, when more stringent containment measures were introduced in the expectation of the risk of increased social interactions during the Easter holiday.

**Figure 5.**
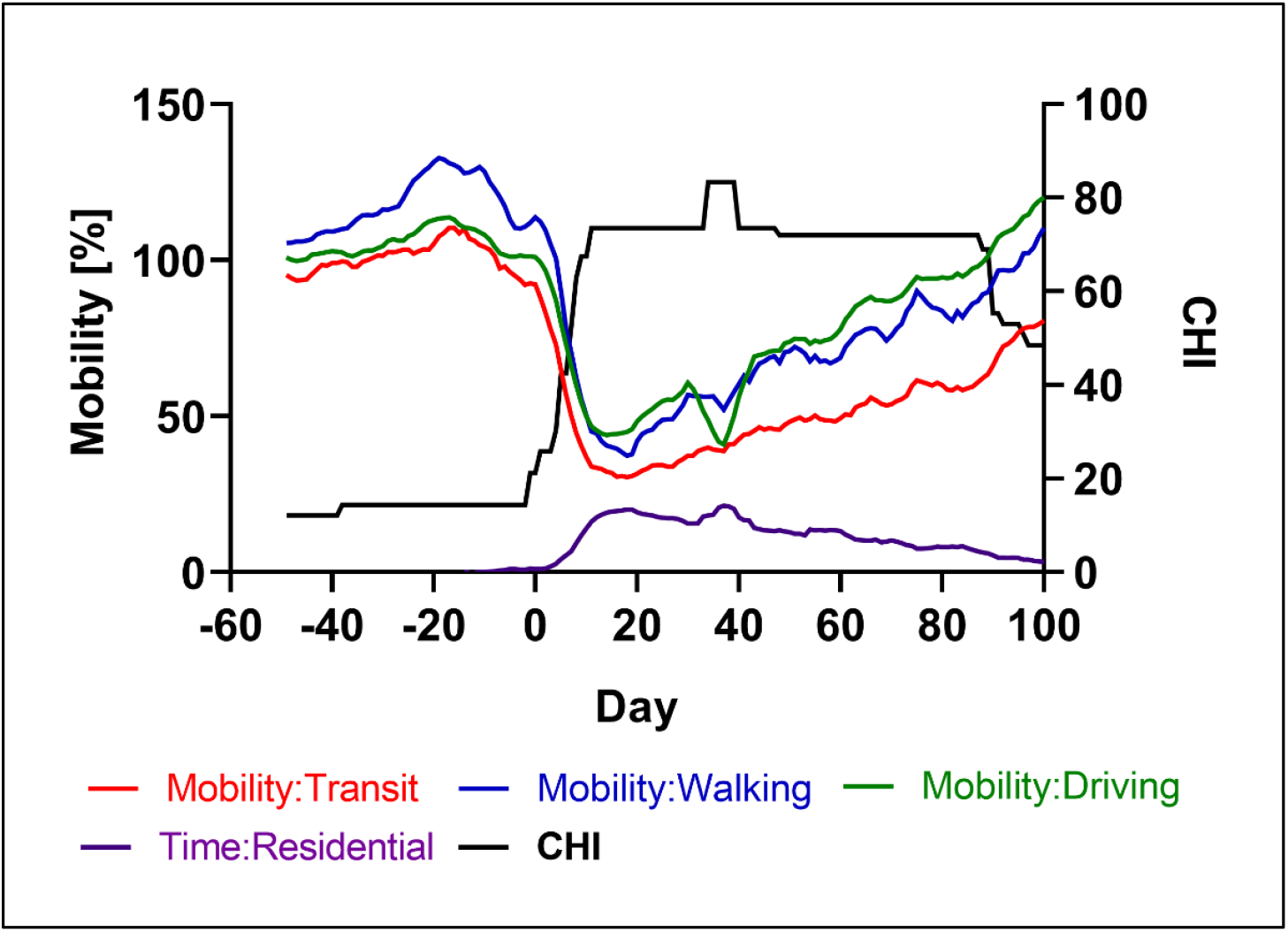
Mobility and the Containment and Health Index (CHI) in the Slovak Republic. Vertical axis: change in mobility relative to the baseline (for details, see Datasets and Methods section). Horizontal axis: day relative to the start of epidemic (day 1= 6 March 2020).

### 3.5 European countries displayed similar patterns of mobility and stringency of the containment measures over calendar days, but these patterns differed relative to the beginning of their epidemics

Interestingly, European countries with the lowest (SK, BG, HU) and highest (LU, BE, ES) cumulative incidences in the first pandemic wave displayed similar calendar dates of mobility changes. Specifically, Slovakia, Bulgaria and Hungary experienced increasing mobility from January 13^th^ up to February 18^th^-19^th^, but decreasing mobility thereafter, up to March 22^nd^ (Slovakia) or March 27^th^ (Bulgaria and Hungary), and the same time courses were also found for Luxembourg and Belgium (Supplemental figures 9-14). In contrast, however, the first COVID-19 cases were diagnosed later in the three lowest incidence countries than in the three highest incidence countries (Table 1). This finding indicates consistent patterns of mobility when considered relative to the calendar date, but different patterns of mobility with respect to the beginning of epidemics in these countries.

The similar patterns of mobility in calendar periods did not project into the similar disease occurrences across these countries, because the countries differed in their times of disease introduction. Decreased mobility coincided in time with disease introduction in the lowest incidence countries. In contrast, however, the highest incidence countries showed delayed reduction in mobility relative to first days of their epidemics (Table 4). For instance, reduction of driving and walking mobility below 50% of the baseline took 8 days for Slovakia, but 43 days for Belgium counting from the day of first case diagnosis. Likewise, on the day of the first case diagnosis, mobility has been already decreasing in Slovakia, Bulgaria and Hungary, but it has been still increasing or not changing in Belgium, Spain and Luxembourg relative to the baseline. Thus, timing of mobility changes, although similar across the six countries, was more favourable in the lowest incidence countries, if assessed relative to the time of the first confirmed disease occurrence (Figure 6).

**Table 4.** Containment Health Indices (CHI) reached at day 1, 7 and 14 from the first diagnosed cases and maximum achieved CHI in three European countries with highest and lowest cumulative incidences of COVID-19 for the first epidemic wave.

|  | Containment Health Index (CHI) |  |  |  | Time from the 1<sup>st</sup> diagnosed case to the reduction of mobility below 50% of the mobility on 13-01-2020 (in days) |  |  |
| --- | --- | --- | --- | --- | --- | --- | --- |
|  | Day 1 | Day 7 | Day 14 | Maximum | Driving | Walking | Transit |
| Slovakia | 25.76 | 52.27 | 73.48 | 83.33 | 8 | 8 | 7 |
| Bulgaria | 20.45 | 44.7 | 62.88 | 71.97 | 8 | 8 | - |
| Hungary | 21.21 | 28.03 | 67.42 | 75.00 | 16 | 14 | - |
| Luxembourg | 0 | 24.24 | 46.97 | 77.27 | 16 | 17 | 15 |
| Belgium | 9.09 | 9.09 | 9.09 | 77.27 | 43 | 43 | 41 |
| Spain | 21.21 | 21.21 | 21.21 | 80.30 | 43 | 42 | 42 |

**Figure 6.**
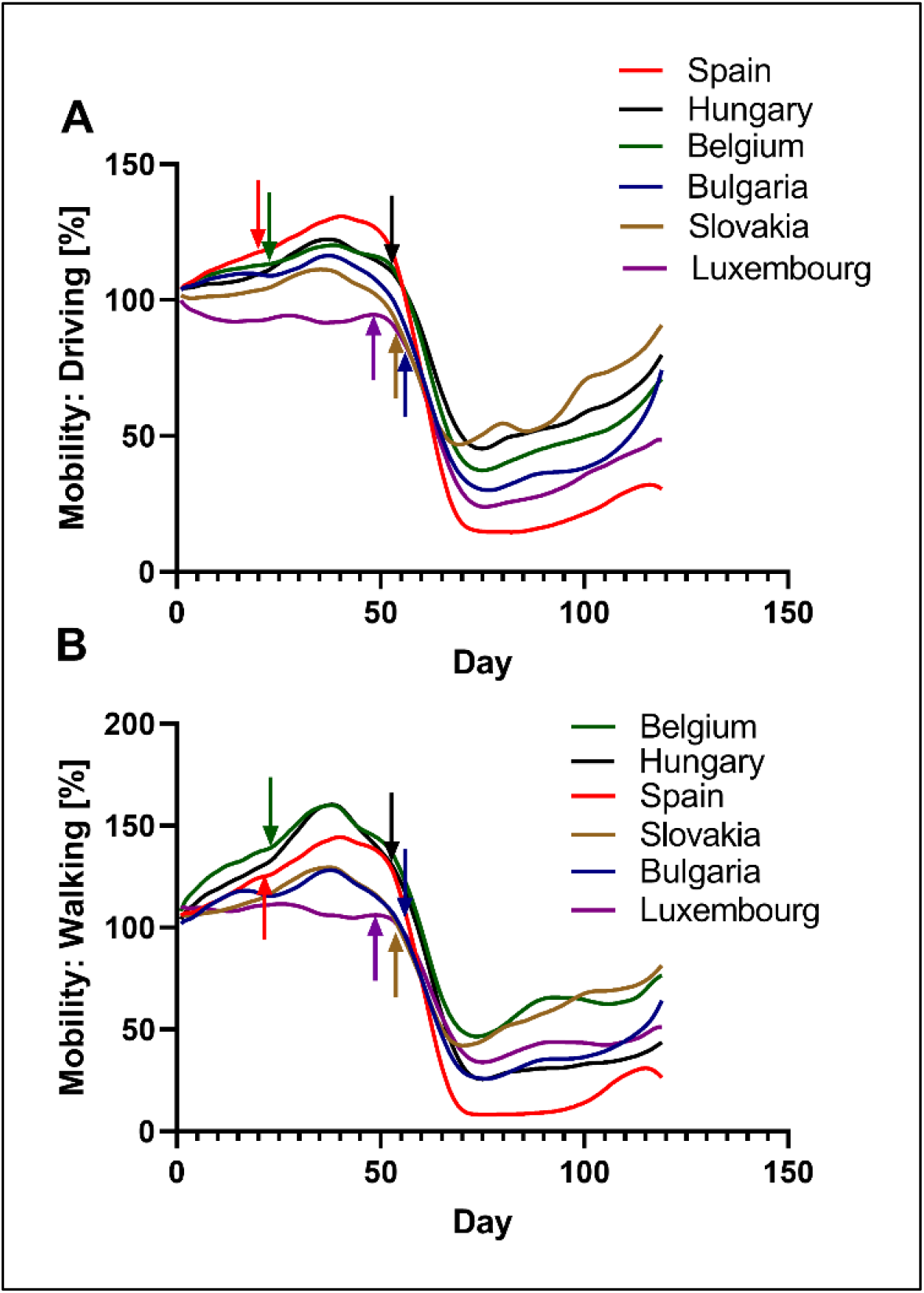
Mobility trends for driving (A) and walking (B) across 3 countries with the highest and 3 countries with the lowest cumulative incidences over the first wave of COVID-19. Color-coded arrows indicate days of the first confirmed COVID-19 cases in each country.

**Figure 7.**
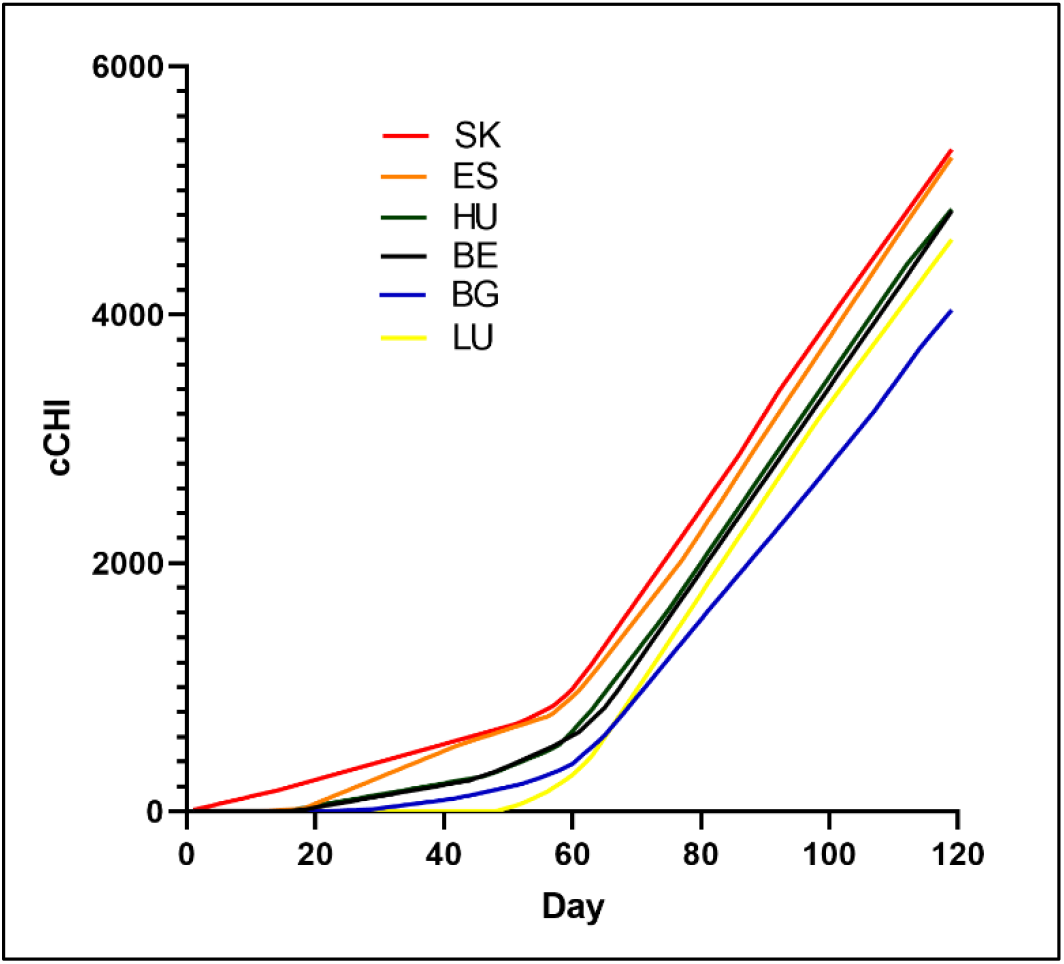
Overall stringency trends by calendar days for 3 countries with the highest (LU, BE, ES) and 3 countries with the lowest (SK, BG, HU) cumulative incidences over the first wave of COVID-19 epidemics. cCHI: cumulative Containment and Health Index accumulated from 13^th^ January 2020. Day: calendar day (1=13^th^ January 2020).

Intriguingly, the overall stringency of containment measures, represented by the cCHI values, was remarkably similar in high-incidence Spain and low-incidence Hungary, when stringency was assessed based on calendar days (Figure 7). Nevertheless, the overall stringency was still higher in Hungary than in Spain, when assessed based on time from the beginning of epidemic (Table 2).

### 3.6 Higher stringency of the containment measures adopted later in the course of epidemics could not fully compensate for delays in initial public health response to COVID-19

In Slovakia, Bulgaria, Hungary and Spain, the CHI values exceeded 20 for the containment measures, which were in place at the day 1 (Table 4). However, unlike SK, BG and HU, whose CHI values raised above 60 at the day 14, Spain displayed no further increase for the next 13 days, and remained at the CHI∼20. In contrast, Luxemburg and Belgium showed CHI<10 at the day 1, and all the three highest prevalence European countries displayed CHI<50 at the day 14. Consequently, lowest incidence countries differed not only with respect to timing of their mobility changes relative to the days of the first disease occurrence, but also in stringency of the containment and health measures reached at days 1 and 14 of their epidemics.

Eventually, the three highest prevalence countries adopted more stringent containment measures than two of the three lowest prevalence countries, in order to control their alarmingly growing epidemics. Nevertheless, this delayed ramping-up of the containment measures did not help to avoid high disease occurrences in Luxembourg, Belgium and Spain during their first epidemic waves (Table 4).

## Discussion

Pandemic of COVID-19 stressed the urgency of ongoing discussions in the field regarding the effect of non-pharmaceutical interventions (NPIs) on the spread of epidemic disease. Various NPIs adopted in response to the past pandemics of respiratory viral diseases have reportedly demonstrated effectiveness, if used in their combinations, and if adopted with appropriate timing and duration, even though disease transmission tended to reoccur when the NPIs were later relaxed^16^.

For instance, during the 1918-1919 influenza pandemic, most commonly adopted NPIs in the USA involved a combination of school closures with public gathering bans, adopted for median duration of 4 weeks. Early and sustained adoption of these NPIs was reportedly associated with decreased excess mortality rates from pneumonia and influenza, delayed time to peak mortality, and lower overall mortality^17^. Although no single intervention showed association with improved outcome of the 1918-1919 pandemic influenza, early implementation of the multi-layered NPIs encompassing closure of schools, churches and theaters, was associated with reduced peak mortality rates^16^. More recently, the use of NPIs occurred in 2002-2003 during SARS^18,19^, and in 2009-2010 during the H1N1pmd09 influenza pandemics^20^.

Effectiveness of the NPIs have been questioned within scientific and professional community regarding the strength of existing evidence^21^, but also in the general public, which is vulnerable to the COVID-19 related misinformation and conspiracy beliefs about the reasons behind the NPIs, and about their alleged harm and inefficacy^22^.

Our results indicate that higher stringency of containment measures accumulated over days preceding the day of first diagnosis of COVID-19 is associated with lower cumulative incidence of confirmed cases over the epidemic wave. The same finding holds for cumulative stringency of containment measures adopted over first 7 or more days of epidemic. Our finding supports the effectiveness of early adopted and stringent containment measures in reducing the overall number of COVID-19 cases. This finding is consistent with expectations derived from the epidemiological theory, which posits that the reduction of person-to-person contact rates through containment measures leads to the reduction of reproduction number, which in turn results in the lower total number of infections, as well as the longer time to the peak daily incidence, and the lower peak of daily incidence^23^.

Our study is subject to some potential limitations that need to be considered to allow an objective assessment of our results. Firstly, our estimation of last days of the first pandemic waves in some countries was affected by complexity of epidemic curves and their first and second order derivatives. As a result, we may have underestimated duration of the first epidemic waves and cumulative incidences in some countries, such as Sweden and Poland. Nevertheless, if we consider alternative durations of first epidemic waves of 137 days and 168 days for Sweden and Poland, respectively, our findings of associations between the cumulative stringency of containment and cumulative incidences would not change.

Secondly, cumulative pre-epidemic CHI values were not weighed with respect to their distance from the day 1 of epidemics, which means that stringent measures adopted for a short time and lifted long before the day 1 would count the same in the cCHI as stringent measures adopted closer to the day 1 and still in place at the beginning of epidemics. Indeed, the 28 European countries differed in (i) the length of pre-epidemic periods counted arbitrarily from the 1^st^ January 2020 to the day preceding day1 of their epidemics, and (ii) times when they adopted their first containment measures (Table 1). This in turn allowed individual countries to accumulate CHIs over different times. For example, France accumulated pre-epidemic CHIs over just 1 day, Finland over 2 days, and Belgium over 6 days, while Bulgaria and Slovakia accumulated pre-epidemic CHIs over 34 and 65 days, respectively. Nevertheless, we have not identified any country among considered 28 European countries that would adopt some containment measures in pre-pandemic period after 1^st^ January 2020 only transiently and lift them before the day 1 of epidemic. As a result, the potential bias in determining pre-epidemic cumulative CHIs is likely of limited influence.

Lastly, reduction of mobility was only considered relative to the baseline mobility in the same country at a specified period of time, which does not permit comparison of absolute mobilities among different countries. For instance, the number of registered passenger cars in two countries with similar areas such as, Denmark and Estonia can be considerably different (2,329,580 vs. 653,000 in 2014)^24^ in spite of their similar geographic areas, which implies different absolute mobilities and different effect of the relative reduction of mobility expressed as percent of baseline values. This limitation does not affect comparison across countries with respect of times corresponding to changes in patterns of mobility.

Early adoption and stringent containment measures were previously found to be associated with lower number of cases and deaths due to COVID-19. For instance, implementation of restrictions on gatherings and public events in New Zealand when number of daily cases was in single digits reduced the number of COVID-19 deaths by at least ten times relative to the number of deaths expected in the absence of stringent containment measures^25^. Likewise, cities in China that pre-emptively suspended intra-city public transport and banned public gatherings and entertainment venues had in average 33.3% fewer cases during the first weeks of their outbreaks, compared with cities that started control later^26^. It should be also noted here that the Wuhan city shutdown and cordon sanitaire imposed on 23^rd^ January 2020 was critical for the suppression of epidemic in China, together with the NPIs adopted in other cities. This can be demonstrated by comparison of the number of cases in China outside Wuhan by the day 50 of the epidemic (29,839 cases) and predicted estimates for the number of cases for the different scenarios: (i) without the cordon sanitaire around Wuhan but with the NPIs in other cities: 199,000 cases; (ii) with cordon sanitaire, but without the NPIs in other cities: 202,000 cases; and (iii) without any intervention: 744,000 cases^26^.

The need for early adoption of containment measures was also demonstrated by Loewenthal et al. who found strong correlation between the time at which containment measures were initiated and the number of deaths^27^. Based on their results, delay of 7.49 days in initiating containment measures would result in two-fold increase in the number of deaths. Interestingly, this study indicated that the response time was more important than its strictness.

Lastly, a multi-method study of efficacy of individual NPIs implemented in March-April 2020 on 79 territories found that cancellation of small gatherings (closure of shops, restaurants, and gatherings of 50 persons or less), closure of educational institutions and border restrictions were most efficient in reducing effective reproduction number (Rt) when assessed by four different methods of analysis^28^. Additionally, increased availability of personal protective equipment, individual movement restrictions and national lockdowns were consistently identified as efficient. In contrast, the least effective interventions reportedly included tracing and tracking measures, enhanced capacity for testing and case detection, as well as border health checks and environmental cleaning^28^.

Our finding of inverse correlation between cCHI values and cumulative incidences of COVID-19 is strengthened by controlling for population density. This is in line with previously reported finding that higher population density renders social distancing more difficult and that the effect of containment measures have been stronger in countries with lower population densities^25^. Further, our finding of statistically significant and positive correlation between population density and cumulative incidence of COVID-19 across 28 European countries agrees with previous report on greater rates of transmission of SARS-CoV-2 in the U.S. counties with greater population densities^29^. On the other hand, our finding differs from apparent no association between population density and number of COVID-19 cases per 100,000 people across numerous global locations, which was reported by other investigators^30^. This apparent lack of association, which was used as an argument to support numerous benefits of dense, mixed-use neighbourhoods even in pandemic times, was likely caused by the third-variable effect of the containment stringency on the disease occurrence. As a result, hyper-dense metropolitan areas Singapore, Hong Kong, Tokyo and Soul with stringent NPIs in place could still reach low cumulative incidences, in spite of their high population densities.

The resistance of governments to early adoption of the NPIs stems mostly from their potential for negative economic and social consequences^31^. Indeed, the COVID-19 pandemic have been considered by the World Trade Organization (WTO) and Organization for Economic Cooperation and Development (OECD) as the largest threat to global economy since the global financial crisis of 2008-2009^32^. Nevertheless, the negative impact of NPIs on economic activity has been challenged. For instance, a recent study by Correia et al. implied that economic disruptions during the 1918-1919 influenza pandemic in the USA were attributable to the impact of pandemic rather than to the impact of the associated public health responses^33^. Moreover, this study demonstrated that the US cities that responded earlier and more aggressively to the 1918-1919 pandemic did not experience worse economic disruptions than cities that adopted lenient NPIs later. In contrast, they reportedly tended to grow faster after the pandemic was over^33^.

The resistance of government agencies towards the adoption of NPIs, or their re-adoption in the successive epidemic waves, can also reflect evolving public mistrust and unwillingness to comply with the NPIs, which can lead to overturning expert recommendations when pressed by public opinion.^34^ Therefore, the purpose of this study was to assess the effectiveness of containment measures to mitigate or suppress the spread of COVID-19 in order to inform future response to COVID-19. We can reasonably anticipate that some of these measures will be necessary in the future, at least locally, even if the effective and safe vaccines become available to the general public. The appropriate NPIs will need to be adopted swiftly in locations where contact rates and vaccination coverages would allow disease transmission in the population.

Intriguingly, top three countries with lowest cumulative incidences of COVID-19 over the first epidemic waves received relatively poor ranking among 195 countries by the Global Health Security Index (GHS) in the category “Rapid Response”. The GHS Index evaluates capabilities of 195 countries to prevent, detect, and rapidly respond to public health emergencies, and its “Rapid Response” category specifically evaluates “rapid response to and mitigation of the spread of an epidemic”^35^. The scores and ranks assigned to these countries were as follows: Hungary (score: 52.2%, rank: 33), Slovakia (score: 34.1%, rank: 105), and Bulgaria (score: 21.7%, rank: 170). In contrast, two of the three countries with highest cumulative incidences of COVID-19 were ranked more favorably by the GHS: Belgium (score: 47.35%, rank: 53) and Spain (score: 61.9%, rank: 15), while Luxembourg (score: 27.3%, rank: 139) was not ranked as favorably as Belgium and Spain, but still better than Bulgaria. Our findings question the value of the GHS scoring at least in this context, which is consistent with conclusion of other investigators who found poor predictive performance of the GHS scoring of the OECD countries in the context of COVID-19 pandemic^36^.

## Conclusions

Our results support effectiveness of non-pharmaceutical interventions, and specifically selected containment and health measures, in mitigation or suppression of COVID-19 epidemics. Early adoption of stringent containment measures, which lead to the high cumulative CHI values in pre-epidemic periods, together with ramping up of containment stringency during early days of epidemic is associated with lower cumulative incidence of COVID-19 cases. Late adoption of stringent containment measures does not fully compensate for the lack of early response.

## Data Availability

This study only used public datasets, which are identified in the Datasets and Methods section.

https://github.com/CSSEGISandData/COVID-19/find/master

https://github.com/OxCGRT/covid-policy-tracker

## Acknowledgement

Authors thank Dr. Roger Wartell for reviewing this manuscript and providing insightful comments, and Mr. Augustín Rosa for his help with visualisations.

## Supplemental Figures

**Supplemental figure 1.**
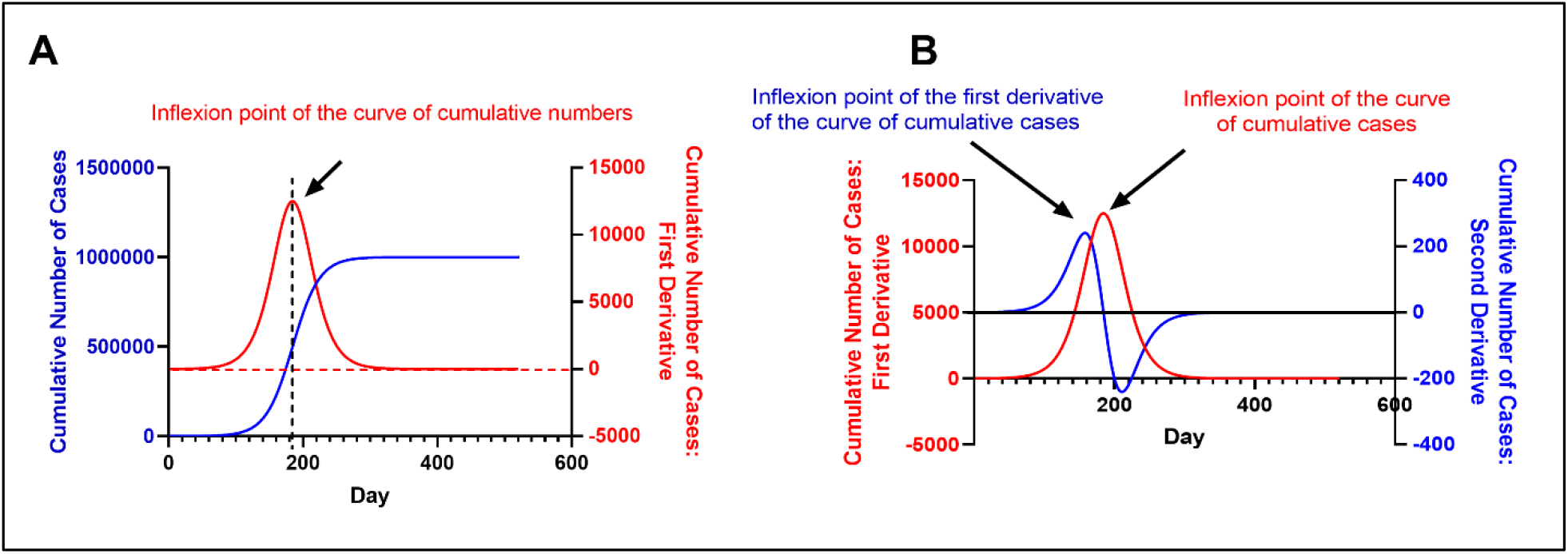
Demonstration of the shapes of the curves for cumulative numbers of cases and their first order (A, B) and second order (B) derivatives. Plotted for classical logistic growth (dN/dt=kN(1-N/K)), using 100 initial cases (N_0_), growth rate of 0.05 day^-1^ (k) and carrying capacity of 1,000,000 (K).

**Supplemental figure 2.**
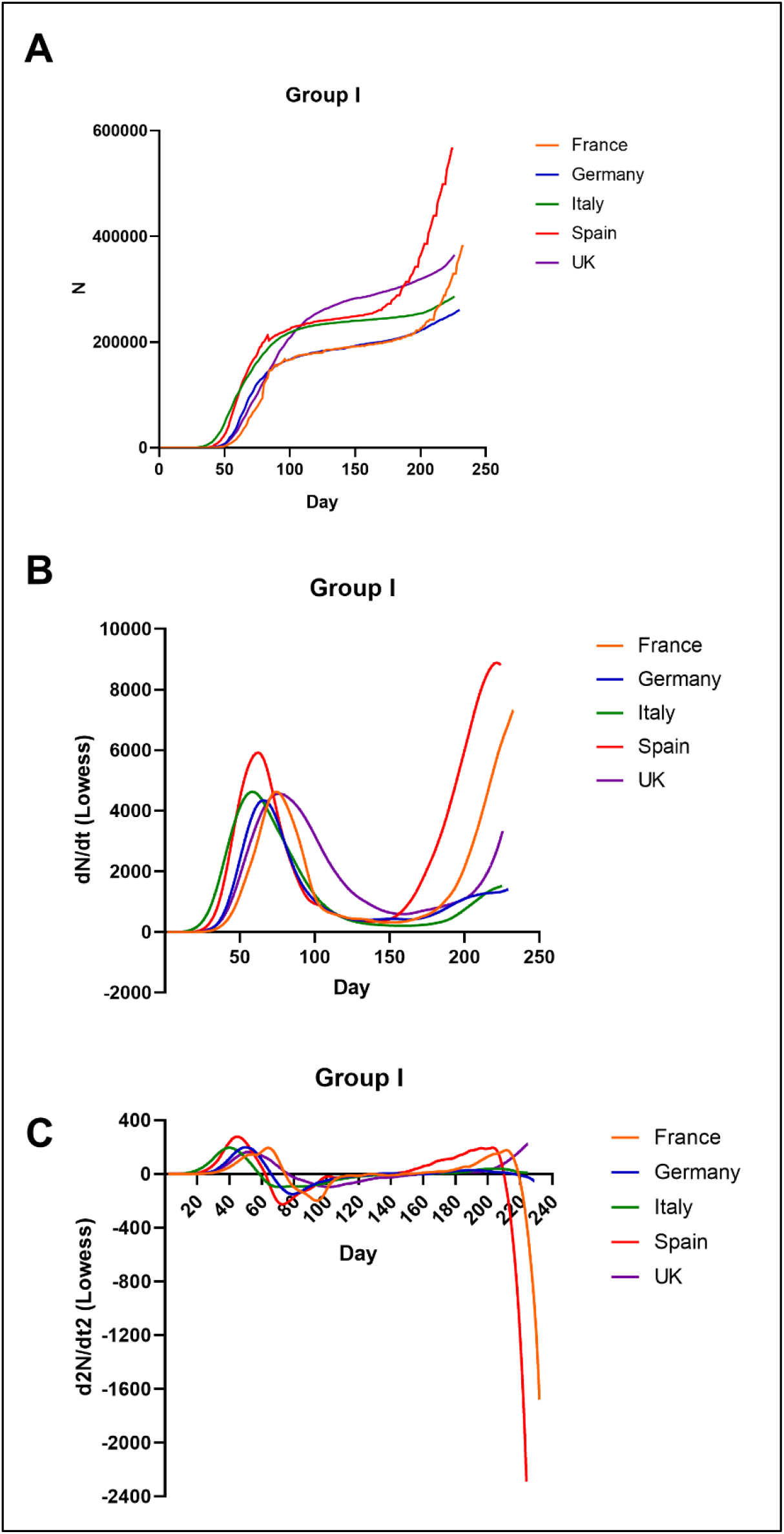
Cumulative number of confirmed COVID-19 cases vs. day since the first confirmed case (A), and its first order derivative (B) and second order derivative curves. Countries in figures 2-6 were grouped based on similarity of cumulative incidences over first waves of epidemics.

**Supplemental figure 3.**
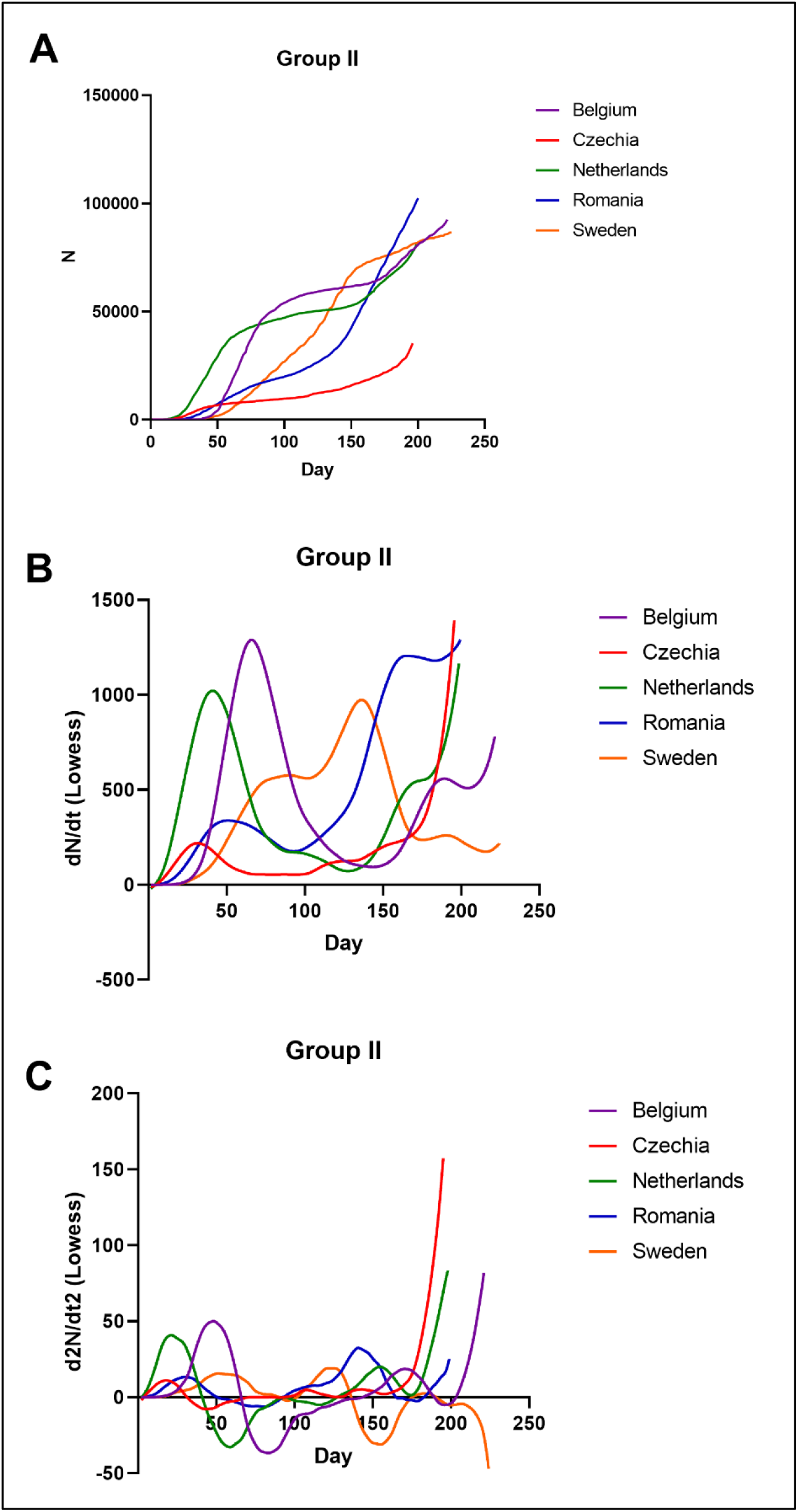
Cumulative number of confirmed COVID-19 cases vs. day since the first confirmed case (A), and its first order derivative (B) and second order derivative curves. Countries in figures 2-6 were grouped based on similarity of cumulative incidences over first waves of epidemics.

**Supplemental figure 4.**
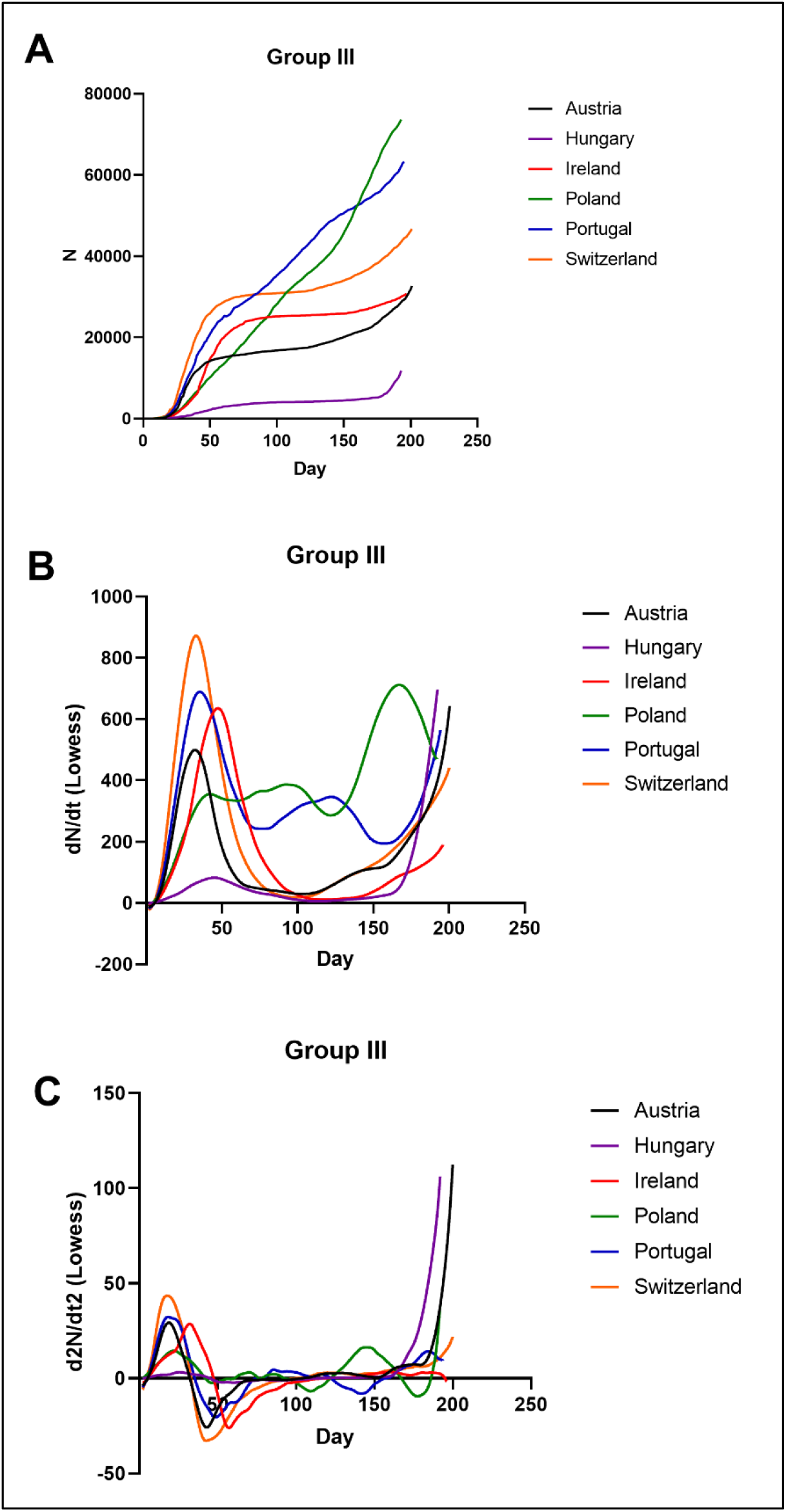
Cumulative number of confirmed COVID-19 cases vs. day since the first confirmed case (A), and its first order derivative (B) and second order derivative curves. Countries in figures 2-6 were grouped based on similarity of cumulative incidences over first waves of epidemics.

**Supplemental figure 5.**
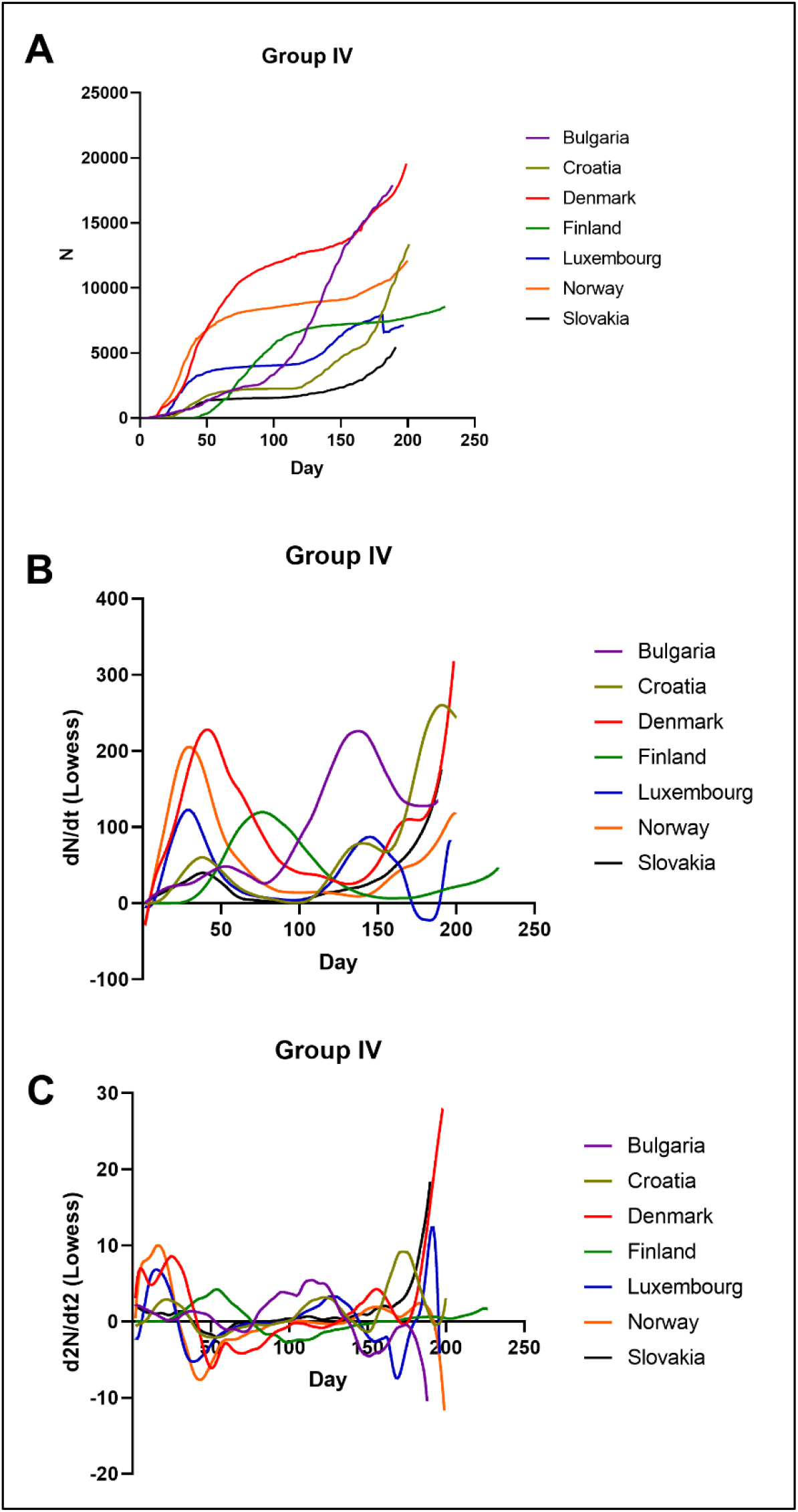
Cumulative number of confirmed COVID-19 cases vs. day since the first confirmed case (A), and its first order derivative (B) and second order derivative curves. Countries in figures 2-6 were grouped based on similarity of cumulative incidences over first waves of epidemics.

**Supplemental figure 6.**
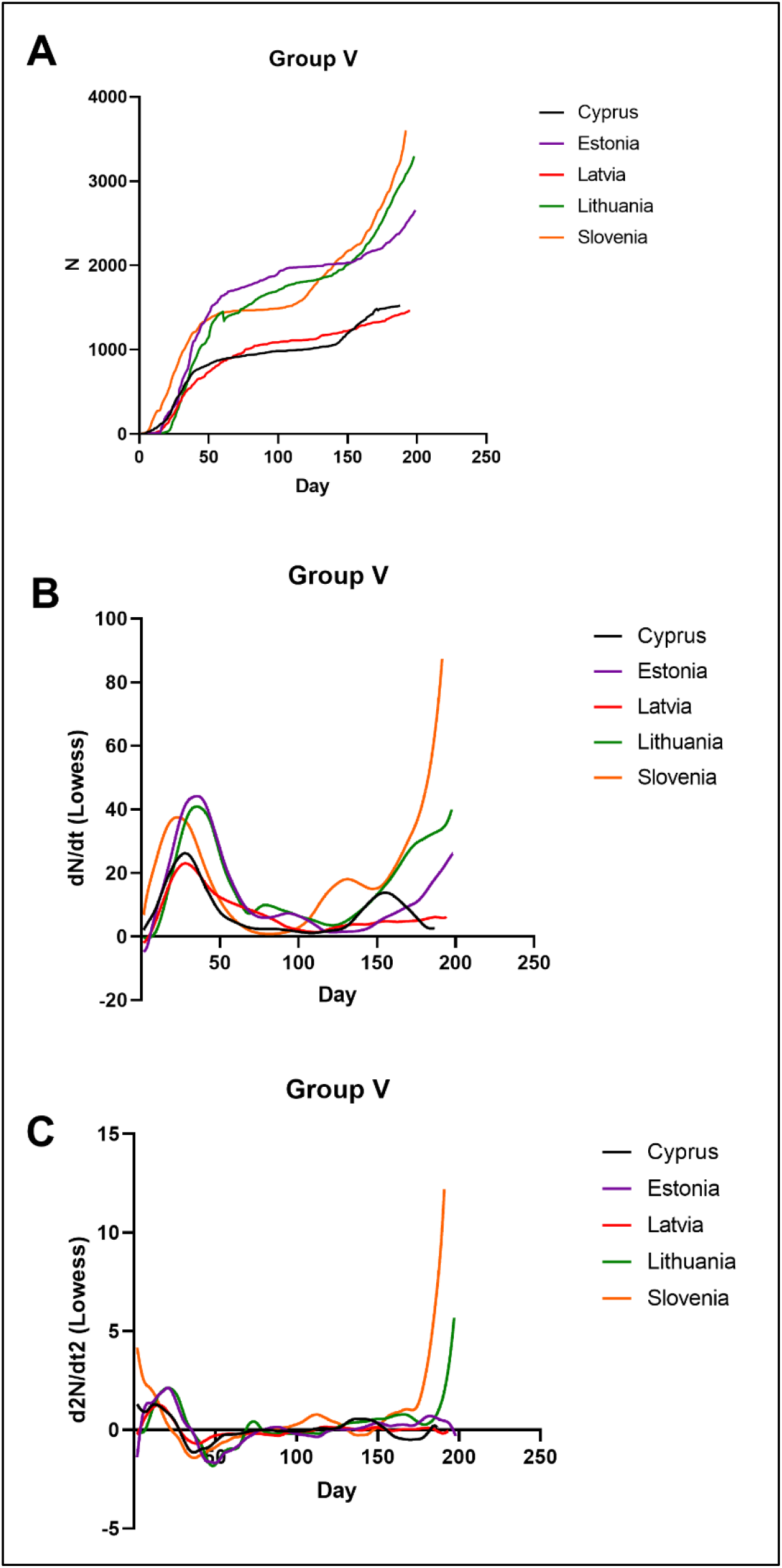
Cumulative number of confirmed COVID-19 cases vs. day since the first confirmed case (A), and its first order derivative (B) and second order derivative curves. Countries in figures 2-6 were grouped based on similarity of cumulative incidences over first waves of epidemics.

**Supplemental figure 7:**
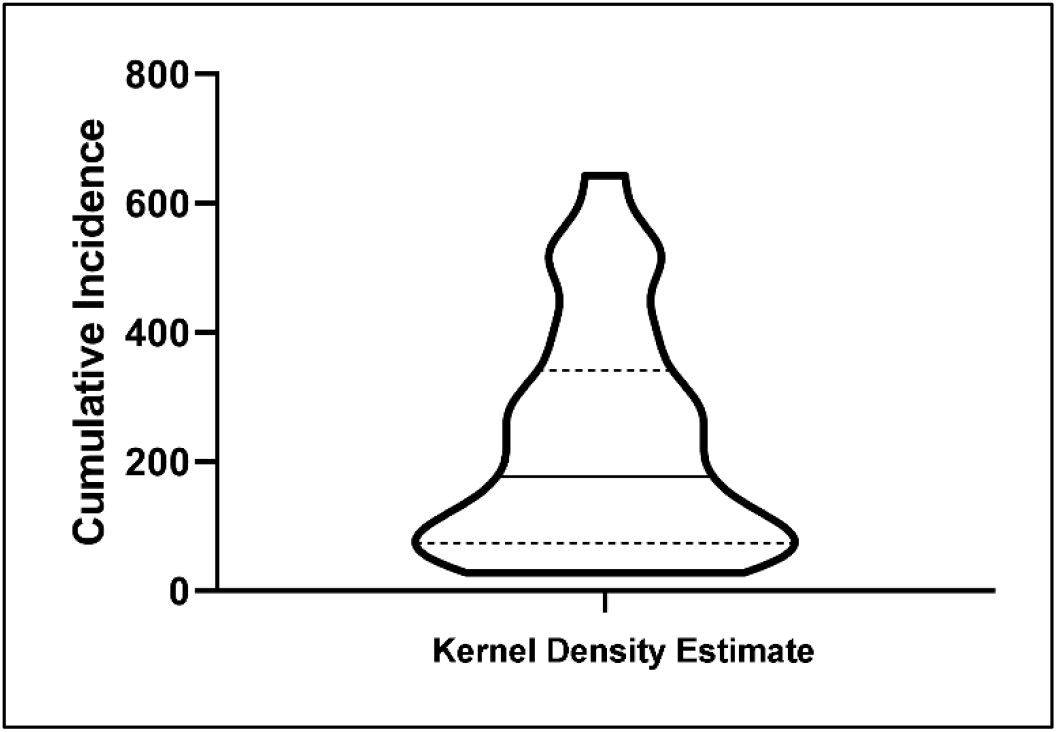
Violin plot for cumulative incidences reached 28 European countries during the first epidemic wave of COVID-19. Dashed lines: quartiles; full line: median.

**Supplemental figure 8:**
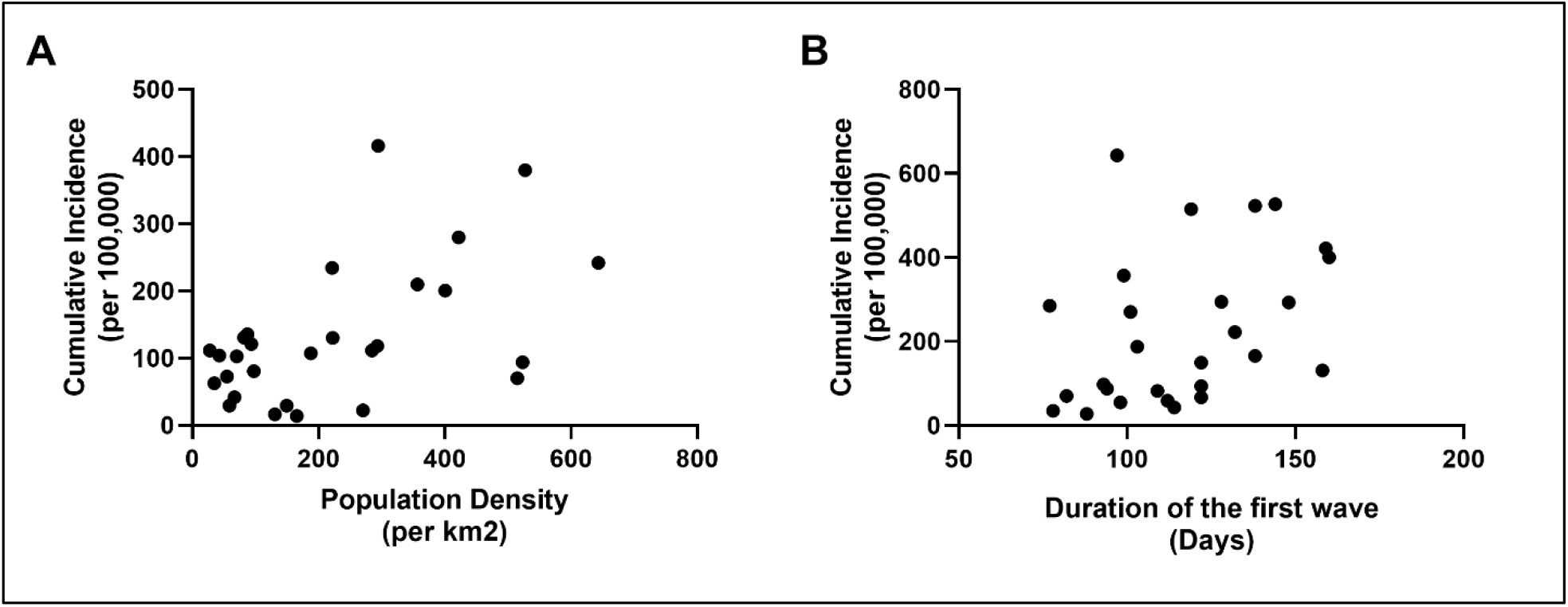
Scatter plots of cumulative incidences reached during the first epidemic waves of COVID-19 in 28 European countries vs population density (A) or duration of the first epidemic wave (B).

**Supplemental figure 9:**
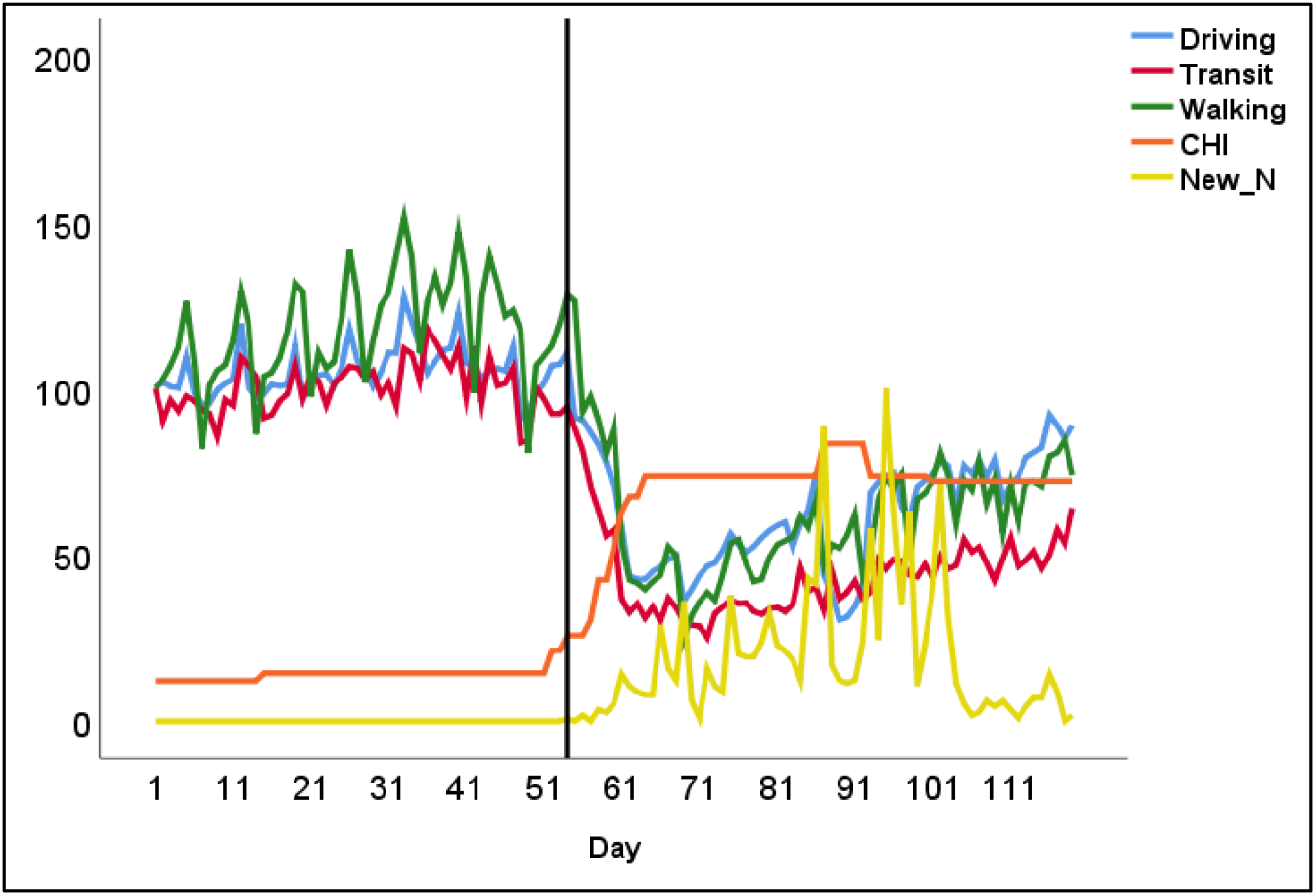
Slovakia - Mobility (driving, transit and walking), containment health index (CHI), and numbers of daily COVID-19 cases normalized to maximum during the period from 13 January (day 1) to 10 May 2020 (day 119). Day 1 = 13 January 2020; Vertical line=day of the first case diagnosis

**Supplemental figure 10:**
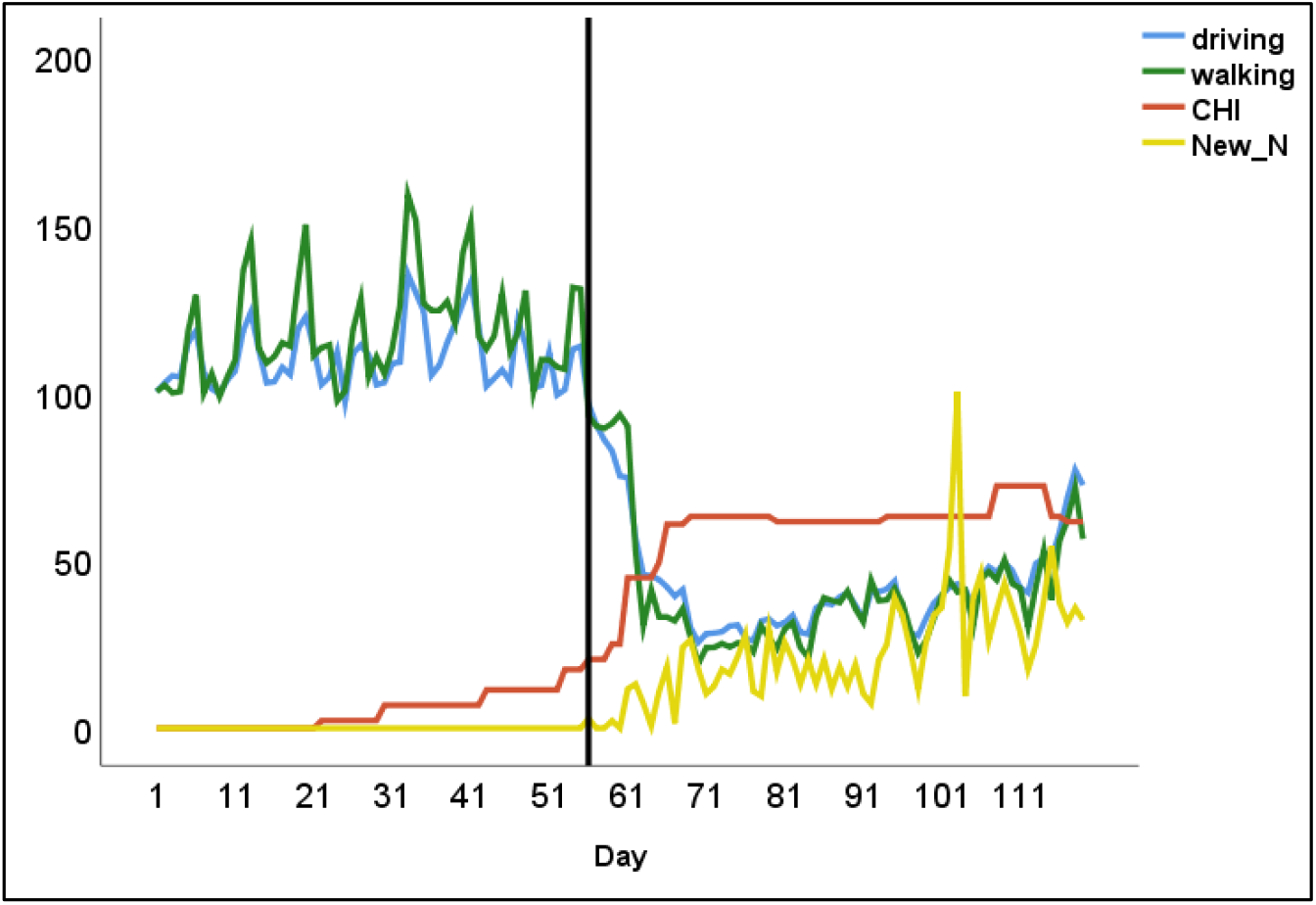
Bulgaria - Mobility (driving, transit and walking), containment health index (CHI), and numbers of daily COVID-19 cases normalized to maximum during the period from 13 January (day 1) to 10 May 2020 (day 119). Day 1 = 13 January 2020; Vertical line=day of the first case diagnosis

**Supplemental figure 11:**
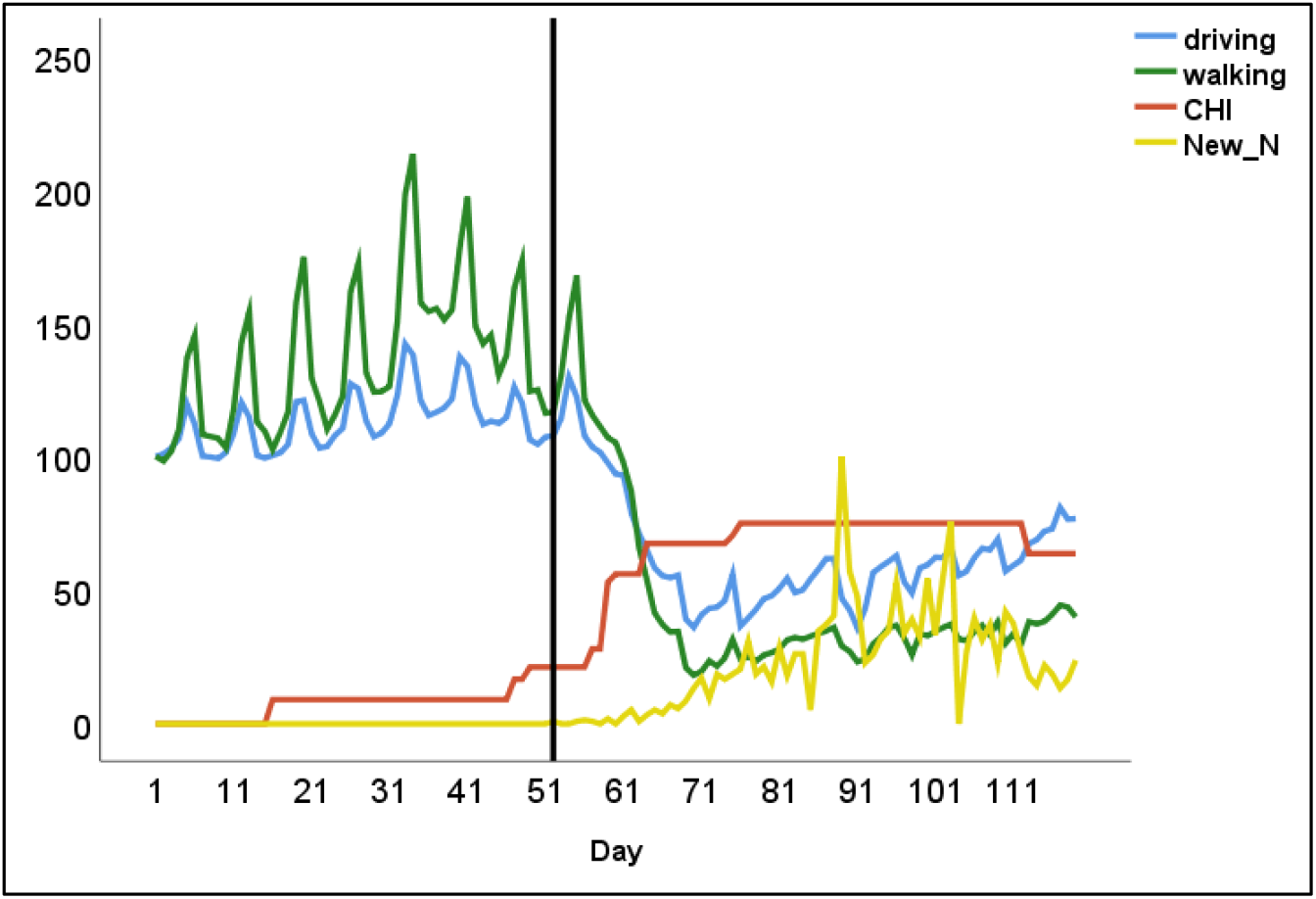
Hungary - Mobility (driving, transit and walking), containment health index (CHI), and numbers of daily COVID-19 cases normalized to maximum during the period from 13 January (day 1) to 10 May 2020 (day 119). Day 1 = 13 January 2020; Vertical line=day of the first case diagnosis

**Supplemental figure 12:**
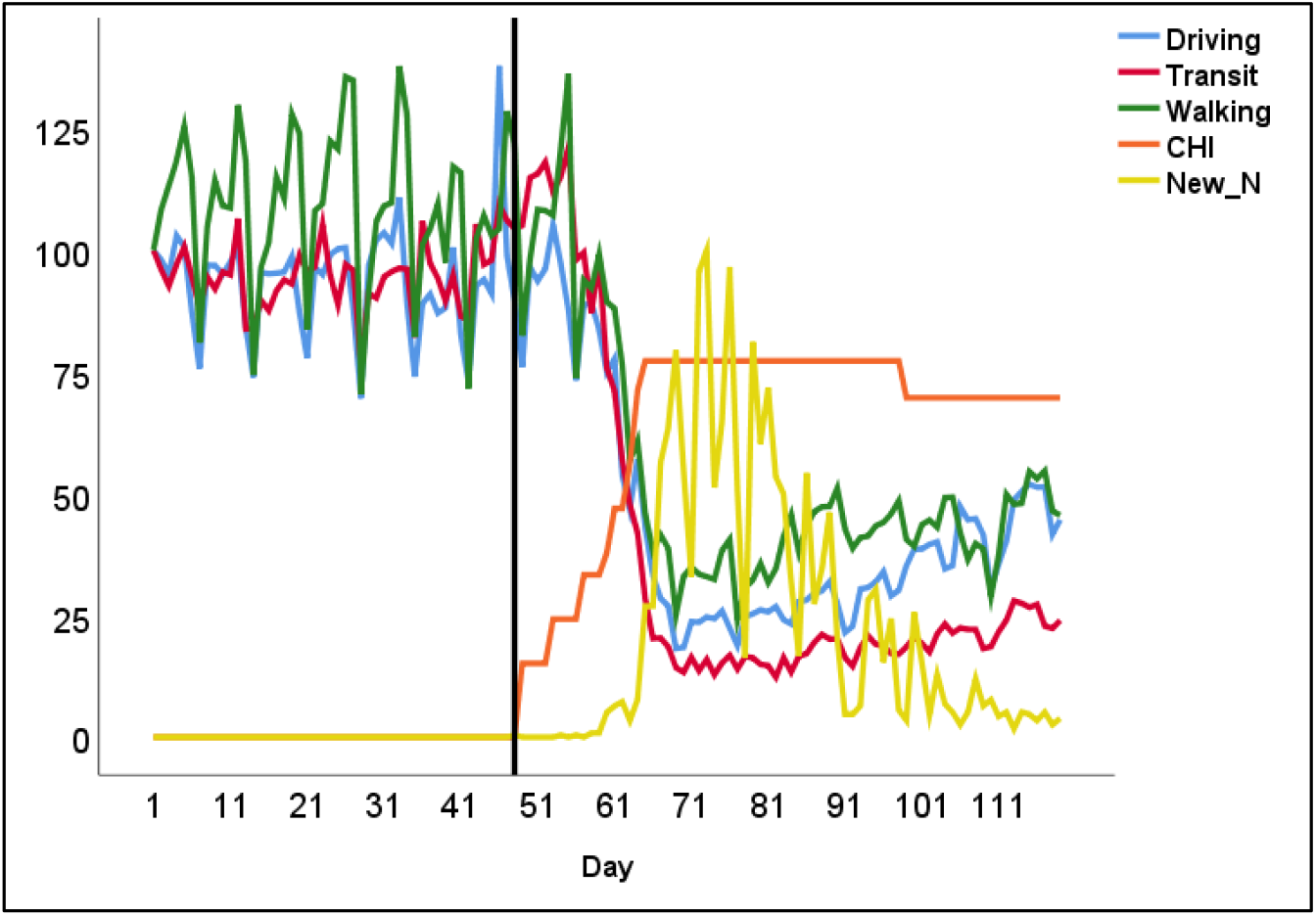
Luxembourg - Mobility (driving, transit and walking), containment health index (CHI), and numbers of daily COVID-19 cases normalized to maximum during the period from 13 January (day 1) to 10 May 2020 (day 119). Day 1 = 13 January 2020; Vertical line=day of the first case diagnosis

**Supplemental figure 13:**
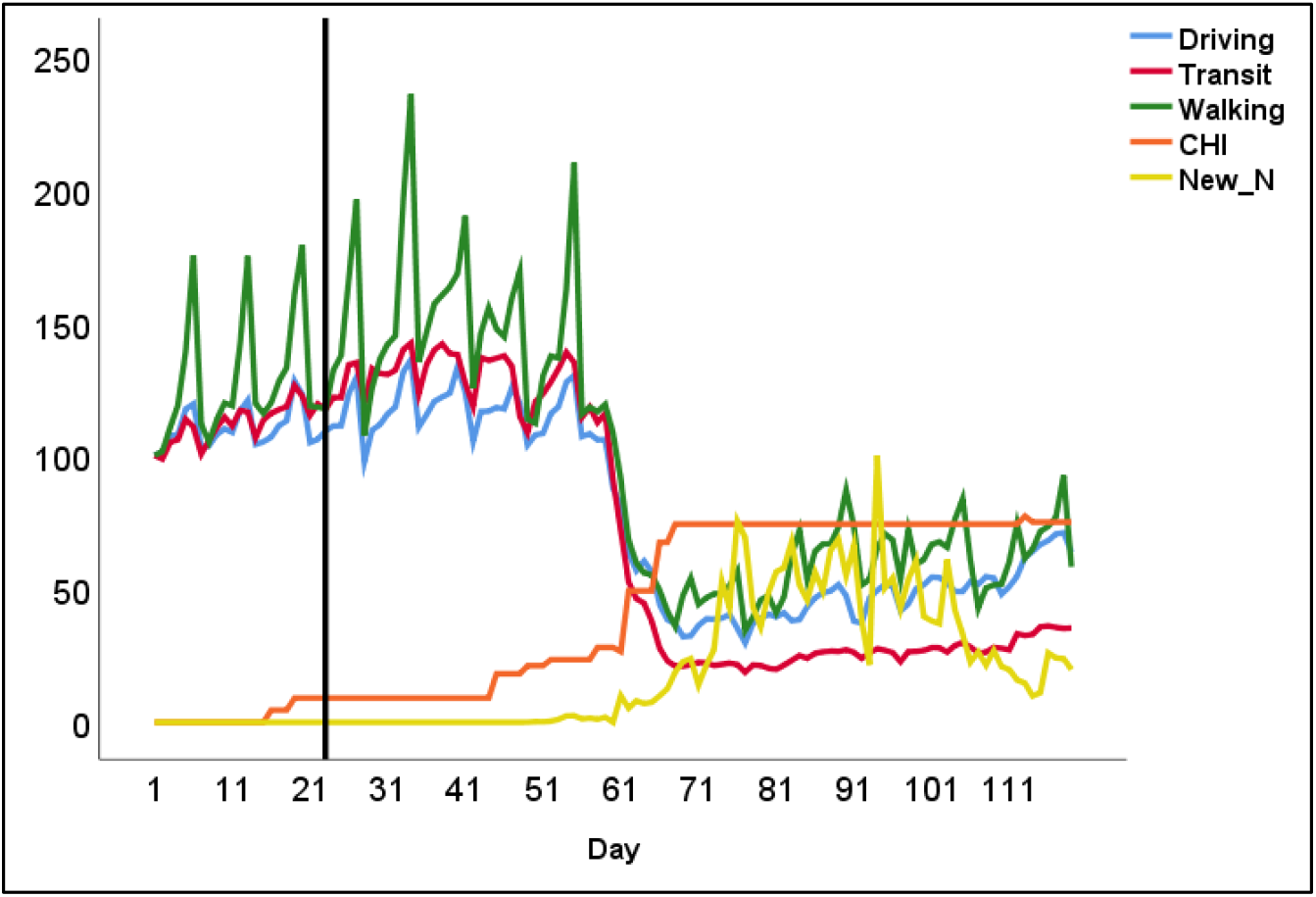
Belgium - Mobility (driving, transit and walking), containment health index (CHI), and numbers of daily COVID-19 cases normalized to maximum during the period from 13 January (day 1) to 10 May 2020 (day 119). Day 1 = 13 January 2020; Vertical line=day of the first case diagnosis

**Supplemental figure 14:**
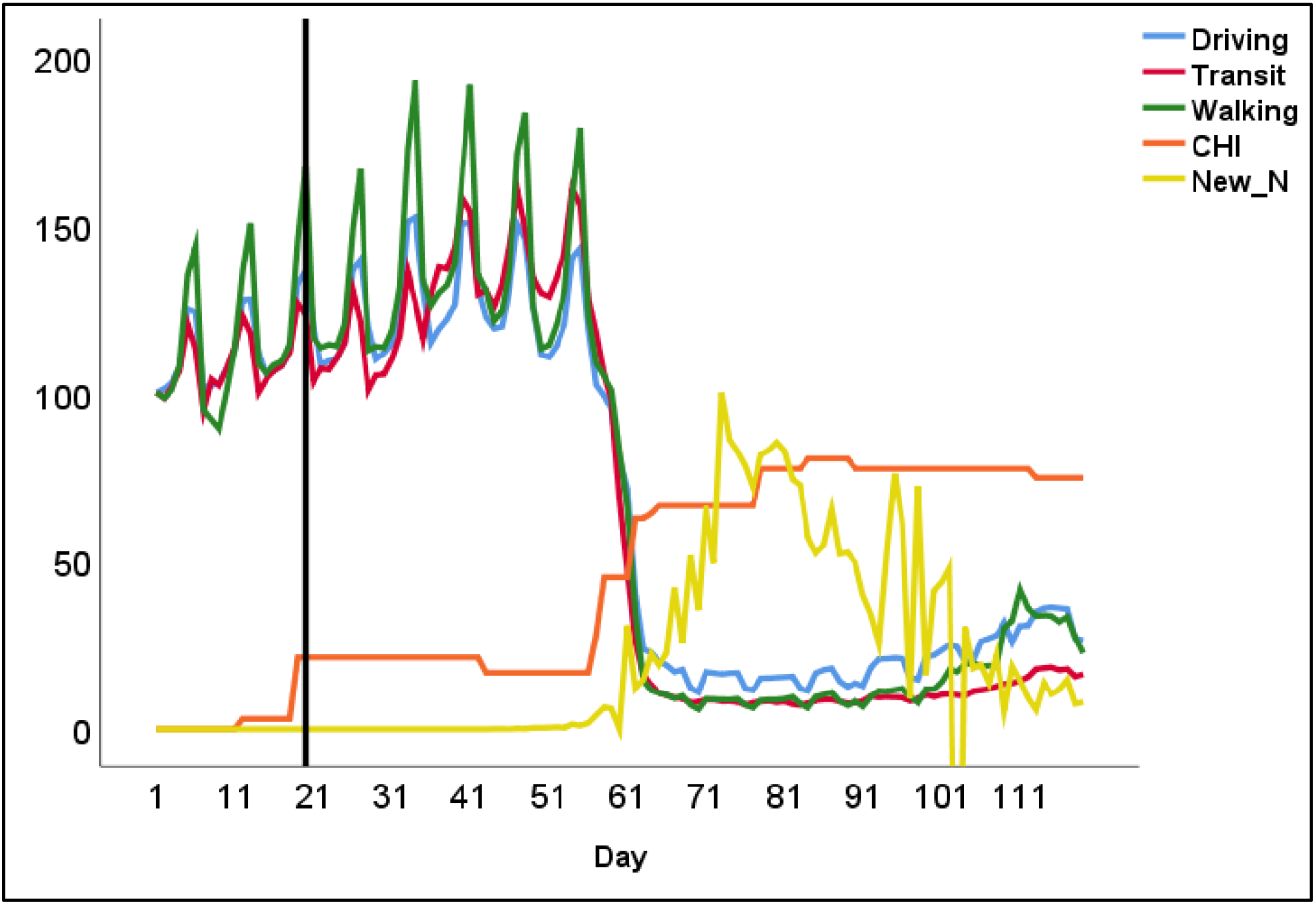
Spain - Mobility (driving, transit and walking), containment health index (CHI), and numbers of daily COVID-19 cases normalized to maximum during the period from 13 January (day 1) to 10 May 2020 (day 119). Day 1 = 13 January 2020; Vertical line=day of the first case diagnosis

